# Malaria-Driven Susceptibility to Invasive Non-Typhoidal *Salmonella*: A Co-infection Study with Implications for Elimination

**DOI:** 10.64898/2026.09.15.26363156

**Authors:** Jonathan D. Sugimoto, Camille Dauvergne, Jean-Paul Kumbukama, Mohamadou Siribie, Marie-France Phoba, Jules Mbuyamba, Seung Eun Kyung, Eunjeong Jeong, Wonchul Chung, Andreia Regina Augusto dos Santos, Rita Soares Barbosa Cardona, Young Ae You, Hyonjin Jeon, Jessica Cowden, Florian Marks, Octavie Lunguya, Birkneh T. Tadesse

## Abstract

**Background:** Invasive non-typhoidal *Salmonella* (iNTS) disease causes high childhood mortality in sub-Saharan Africa. Malaria infection is postulated to increase iNTS risk through impairment of bactericidal immunity. We quantify malaria’s causal contribution to iNTS and explore burden mitigation via integrated antimalarial and iNTS prevention strategies.

**Methods:** We analyzed 443 weeks of surveillance data (September 2017–March 2026) from Kisantu health zone, Democratic Republic of Congo. Analyses included interrupted time series of perennial malaria chemoprophylaxis (PMC; November 2023) and R21/Matrix-M vaccine (October 2024) rollouts, causal mediation analysis, calibration of an age-structured co-infection Susceptible-Infected-Recovered-Susceptible (SIRS) model via the neural ordinary differential equation (ODE) adjoint method, and a 15,435-scenario grid search for NTS infection elimination coverage thresholds.

**Findings:** Malaria burden was 80% lower at PMC full coverage (incidence rate ratio [IRR] 0·197, 95% CI: 0·096–0·401; p<0·001). A declining iNTS trend emerged post-ramp (IRR 0·990/day, 95% CI: 0·981–0·998; p=0·018). The calibrated SIRS model estimated ~12-fold iNTS susceptibility during malaria infection (ψ_M→N_ = 12·0, 95% CI: 3·6–28·3). Combined malaria prevention strategies provided immediate iNTS burden reduction (</= 37%), but full iNTS elimination requires a ≥70% efficacious iNTS vaccine.

**Interpretation:** Malaria control interventions deliver meaningful iNTS co-benefits. This analysis support their prioritization along with further evaluation of integrated malaria-iNTS prevention strategies in high-burden settings. Our results show that full NTS elimination additionally requires a high-efficacy iNTS-specific vaccine.

**Funding:** The Gates Foundation (INV-047715, INV-007844, INV-082265, INV-077454), The European and Developing Countries Clinical Trials Partnership (RIA2017S-2027), and the Swedish Styrelsen för internationellt utvecklingssamarbete (17106)

**Research in Context:** *Evidence before this study:* Clinical and mechanistic studies have established that malaria facilitates iNTS disease, including through haem-driven immune dysfunction, with malaria co-infection present in 40–60% of hospitalized iNTS cases. However, this evidence comes from individual-level clinical studies and does not quantify the population-level causal impact of malaria infection on iNTS disease at scale. No prior study has formally measured whether reducing malaria burden in a population also reduces iNTS burden, and intervention coverage thresholds for co-benefits have not been modelled.

*Added value of this study:* This is the first population-level evidence using a quasi-experimental design to examine the effect of malaria control interventions on iNTS incidence in sub-Saharan Africa. Incorporating a 6-month coverage ramp-up and precipitation adjustment, we document an 80% malaria reduction at full PMC coverage (IRR 0·197, 95% CI 0·096–0·401; p<0·001). Seasonal adjustment eliminated the apparent iNTS incidence spike at PMC rollout (fully unadjusted IRR 2·766, 95% CI 1·453–5·267, p=0·002; adjusted IRR 0·972, 95% CI 0·418–2·262, p=0·948), revealing a seasonal confounding artefact, and uncovered a significant declining iNTS trend post-ramp (IRR 0·990/day, 95% CI 0·981–0·998; p=0·018). We calibrate a mechanistic model estimating ~12-fold malaria–iNTS susceptibility enhancement (ψ_M→N_ = 12·0, 95% CI: 3·6–28·3; Mode 1 MAP), subject to identifiability caveats. The elimination boundary analysis shows that a highly efficient iNTS vaccine is necessary for elimination, while combined malaria and iNTS interventions show synergistic effects at sub-elimination coverage.

*Implications of all available evidence:* Malaria control should proceed immediately for multiple co-benefits including iNTS burden reduction. Our modeling results support that integrated malaria–iNTS control strategies offer greater benefits on disease burden than sequential approaches. Future research should confirm these findings in other African settings and measure iNTS burden reduction as R21/Matrix-M is rolled out at scale.

## Introduction

Invasive non-typhoidal *Salmonella* (iNTS) disease – defined as the detection of non-typhoidal *Salmonella* bacteraemia by blood culture – is a leading cause of laboratory-confirmed bacteraemia in sub-Saharan African children,^1, 2^ with case fatality rates of 10–25% even in hospital settings with antimicrobial therapy.^2, 3^ Detected cases represent only a fraction of the true burden; population-level models estimate that most infections are treated empirically or remain undetected entirely.^4, 5^ Despite this mortality burden and the need^6^ and effort^7^ to develop iNTS vaccines, the epidemiological drivers of iNTS disease remain poorly understood at the population level.

A convergent body of clinical and mechanistic evidence suggests that *P. falciparum* malaria actively facilitates iNTS disease.^8, 9^ *P. falciparum*–driven haemolysis releases free haem into the bloodstream; free haem directly inhibits the nicotinamide adenine dinucleotide phosphate (NADPH) oxidase–mediated oxidative killing of *Salmonella* in granulocytes, and haem oxygenase-1 induction impairs bactericidal capacity.^10^ Prospective cohort studies document malaria co-infection in the majority of children hospitalised with iNTS bacteraemia, with population attributable fractions estimated at 40–62%.^2, 11, 12^ Concurrent seasonal peaks of malaria and iNTS in the rainy season are consistent with a causal relationship rather than shared environmental drivers.

Despite this evidence, the population-level magnitude of malaria’s causal contribution to iNTS has not been formally quantified using intervention-based epidemiological methods or calibrated mechanistic models. This gap has policy consequences: if a substantial fraction of iNTS cases is causally attributable to malaria, scaling malaria control interventions could substantially reduce iNTS disease burden even without a licensed iNTS vaccine – and combined malaria–iNTS strategies may be synergistically more effective than either alone.

Two developments make this question newly tractable. Firstly, the licensed R21/Matrix-M (Serum Institute of India, Oxford), a malaria vaccine with 75% phase-3 efficacy in seasonal transmission settings^13^, provides a powerful tool for malaria control in the age group most vulnerable to iNTS disease. Secondly, advances in neural ordinary differential equations (ODEs) enable efficient gradient-based calibration of complex mechanistic models to observed time series, permitting simultaneous estimation of interaction parameters otherwise unidentifiable from aggregate surveillance data.^14^

We present an integrated analysis of weekly malaria and iNTS surveillance data from Kisantu health zone, western Democratic Republic of Congo (DRC), combining quasi-experimental interrupted time series (ITS) analysis, causal mediation analysis, and a calibrated age-structured co-infection Susceptible-Infected-Recovered-Susceptible (SIRS) model to: (1) estimate the population-level causal contribution of malaria to iNTS burden; (2) quantify the malaria-to-NTS susceptibility enhancement coefficient; and (3) identify the combined intervention coverage required to achieve NTS elimination.

## Methods

### Study Setting and Data

The surveillance system methodology is described elsewhere.^15, 16^ In summary, passive surveillance for febrile illness was conducted at healthcare facilities serving residents of Kisantu health zone, western DRC, approximately 100 km southwest of Kinshasa. Blood cultures were obtained from all patients meeting enrolment criteria; concurrent malaria testing was performed using rapid diagnostic test. iNTS disease confirmation was by standard microbiological methods at the Institut National de Recherche Biomédicale (INRB) laboratory located on site in Kisantu. The catchment population was estimated at 168,006 in mid-September 2017, growing at approximately 3% per year, with age structure: <2 years (6·9%), 2–4 years (9·7%), 5–14 years (29·2%), and ≥15 years (54·2%).

We analyzed 443 weeks of weekly data (September 2017–March 2026), focusing on blood-culture confirmed iNTS cases classified into four mutually exclusive categories (malaria only, iNTS only, co-infection, tested-negative) and stratified by four age groups. The study period encompasses the introduction of two anti-malaria interventions: perennial malaria chemoprophylaxis (PMC) consisting of sulfadoxine-pyrimethamine + amodiaquine (introduced on November 1, 2023) and R21/Matrix-M vaccination targeting children under 2 years (introduced on October 1, 2024). Ethics approval: IVI, Institutional Review Board and School of Public Health of Kinshasa, Ethic Review Committee.

### Interrupted Time Series Analysis

We estimated the effect of each intervention on malaria and iNTS incidence using a segmented negative binomial regression with intervention dates as fixed breakpoints:

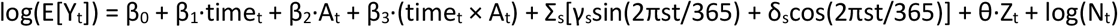

where A_t_ is a linear ramp function (rising from 0 to 1 over the coverage build-up period) to account for gradual rollout of routine immunization delivery, Fourier harmonics (s = 1, 2) control for seasonality, Z_t_ is z-scored weekly precipitation (NASA POWER MERRA2 PRECTOTCORR) to absorb residual rainfall-driven confounding, and log(N_t_) is the log of weekly tested individuals (offset). The slope change term β_3_ accumulates only after the ramp period ends (days since full coverage, not days since introduction). Standard errors were corrected for autocorrelation using Newey-West heteroskedasticity- and autocorrelation-consistent (HAC) covariance^17^, with the Bartlett kernel bandwidth set by the automatic lag-selection rule^18^ (appendix §S6). The primary analysis used a 6-month (183-day) ramp-up; sensitivity analyses tested 3–5 months (appendix table S3). The separate contributions of the coverage ramp, the Fourier harmonics, and the precipitation covariate were assessed by refitting all five nested specifications (appendix table S4).

### Causal Mediation Analysis

We decomposed PMC’s total effect on iNTS disease into its natural direct effect (NDE; not mediated through malaria) and natural indirect effect (NIE; operating through the malaria pathway) using counterfactual mediation analysis within the VanderWeele^19^ potential-outcomes framework, adapted for time-series count outcomes. Negative binomial mediator and outcome models incorporated the intervention indicator, lagged malaria incidence (lags L = 0, 3, 7, 14, 21, 28 days), and Fourier seasonality terms. Parametric g-computation (10,000 Monte Carlo draws) provided estimates of total effect (TE), NDE, NIE, proportion mediated (NIE/TE × 100%), and population attributable fraction; 1,000 bootstrap resamples provided 95% CIs.

### Co-infection SIRS Model

To account for the underlying biological mechanisms driving iNTS susceptibility to malaria – which are not captured through the observational ITS design alone – we developed an age-structured compartmental model for co-circulation of *P. falciparum* malaria (pathogen 0) and NTS (pathogen set 1) across M = 4 age groups, each with susceptible (S_a_), singly infected (I_a,0_, I_a,1_), co-infected (I_a,01_), and recovered/immune (R_a,0_, R_a,1_) compartments. Two interaction mechanisms were modelled.

#### Susceptibility enhancement

ψ_M→N_ ≥ 1 scales the force of NTS infection on malaria-infected individuals, capturing the biologically established mechanism by which haem-driven immune impairment increases NTS susceptibility. A value of ψ_M→N_ = 1 represents no interaction; ψ_M→N_ > 1 indicates facilitation.

#### Post-infection cross-immunity

φ_M→N_ ∈ [0,1] modifies NTS susceptibility in malaria-recovered individuals (initialized at 1·0; no cross-immunity expected a priori between these evolutionarily distant pathogens).

#### Transmission rates followed sinusoidal seasonal forcing

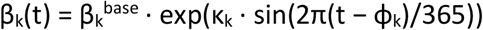, with amplitude κ_k_ and phase φ_k_ estimated during calibration. Weekly precipitation data (NASA POWER MERRA2) were available but excluded from ODE seasonal forcing (Mode 1; a sensitivity calibration Mode 2 included this effect): the sinusoidal term captures endogenous biological transmission cycles, and including an additional precipitation covariate would introduce collinearity without improving identifiability. Seasonal confounding from climate drivers was instead addressed in the ITS model through Fourier harmonics and the precipitation covariate. Observed weekly counts (malaria, iNTS) were modelled as negative binomial with mean equal to model-predicted incidence scaled by reporting fractions (ρ_0_, ρ_1_) and overdispersion parameters (r_0_, r_1_). Full ODE notation is in the appendix.

### Neural ODE Calibration

The ODE was implemented in JAX using the diffrax library^20^ (Dormand-Prince adaptive solver) with gradients computed via the BacksolveAdjoint method, enabling memory-efficient gradient descent through the full time series. We minimized the joint negative log-likelihood of malaria and iNTS case counts using the Adam optimizer with cosine decay (peak learning rate 0·01, 2,000 steps). Parameters were transformed to unconstrained space (log for positive scalars, logit for bounded parameters) and initialized at literature-derived priors. Uncertainty quantification used 100 parametric bootstrap resamples: synthetic counts were drawn from 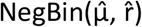 using maximum a posteriori (MAP)estimates, the model was refitted to each replicate, and 2·5th–97·5th percentile CIs were computed. Model identifiability was assessed by profile likelihood analysis for ψ_M→N_.

### Elimination Boundary Simulation

Using the calibrated SIRS model, we projected 5-year iNTS incidence under 15,435 combined intervention scenarios: R21/Matrix-M at 0–100% coverage (5% increments) in children <2 years, with 75% first-year efficacy^13^ and gradual waning; an investigational iNTS vaccine at 0–100% coverage with efficacy swept from 30–90% (seven levels); and PMC at 0%, 25%, 50%, 75%, 100% coverage (83% per-cycle efficacy, four cycles per season^21^). NTS elimination was defined as <1 per million population at year 5, consistent with World Health Organization (WHO) programmatic elimination thresholds for endemic diseases.^22^

### Statistical Analysis

All analyses used Python 3·11. ITS and mediation models used statsmodels 0·14. The SIRS model and training loop used JAX 0·4·25, diffrax 0·5, and optax 0·2. Figures were produced with matplotlib 3·8. Code is available at https://github.com/jsugimot/drc_malaria_ints_ivi.git.

### Role of the funding source

Funders had no role in study design, data collection, analysis, or interpretation.

## Results

### Study Population and Data Summary

Over 443 weeks (September 2017–March 2026), we recorded 12,324 malaria cases, 507 iNTS cases, and 1,316 confirmed co-infections in the Kisantu health zone. The catchment population grew from approximately 168,000 to 216,000 residents during this period. iNTS incidence peaked during rainy seasons (June–September), coinciding with malaria season, consistent with malaria facilitating iNTS disease risk.

### Effect of Anti-Malaria Interventions on Malaria Incidence

Accounting for a 6-month coverage ramp-up and adjusting for precipitation (NASA POWER MERRA2), PMC (November 2023) was associated with an 80% reduction in malaria incidence at full operative capacity (IRR 0·197, 95% CI 0·096–0·401; p<0·001), with no significant post-ramp slope change (IRR 1·002/day, 95% CI 0·998–1·007; p=0·370); full ITS results are in appendix table S1 and visual in Figure 1(a). This estimate was insensitive to the precipitation covariate: removing it, with all else unchanged, gave IRR 0·203 (95% CI 0·100–0·410; p<0·001). The substantially larger reduction relative to a step-function specification (IRR 0·628, 95% CI 0·343–1·151; p=0·132) is therefore attributable to modelling gradual coverage build-up rather than to climate adjustment (appendix table S4).

**Figure 1.**
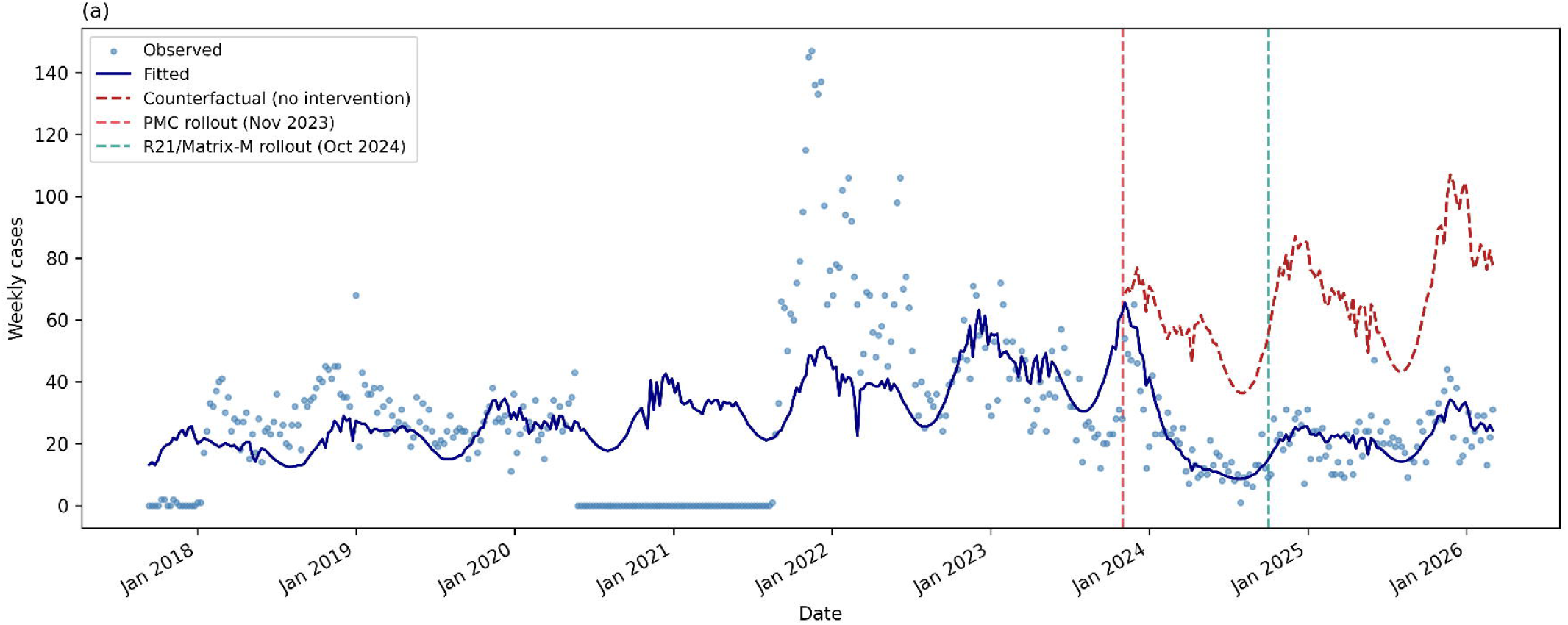
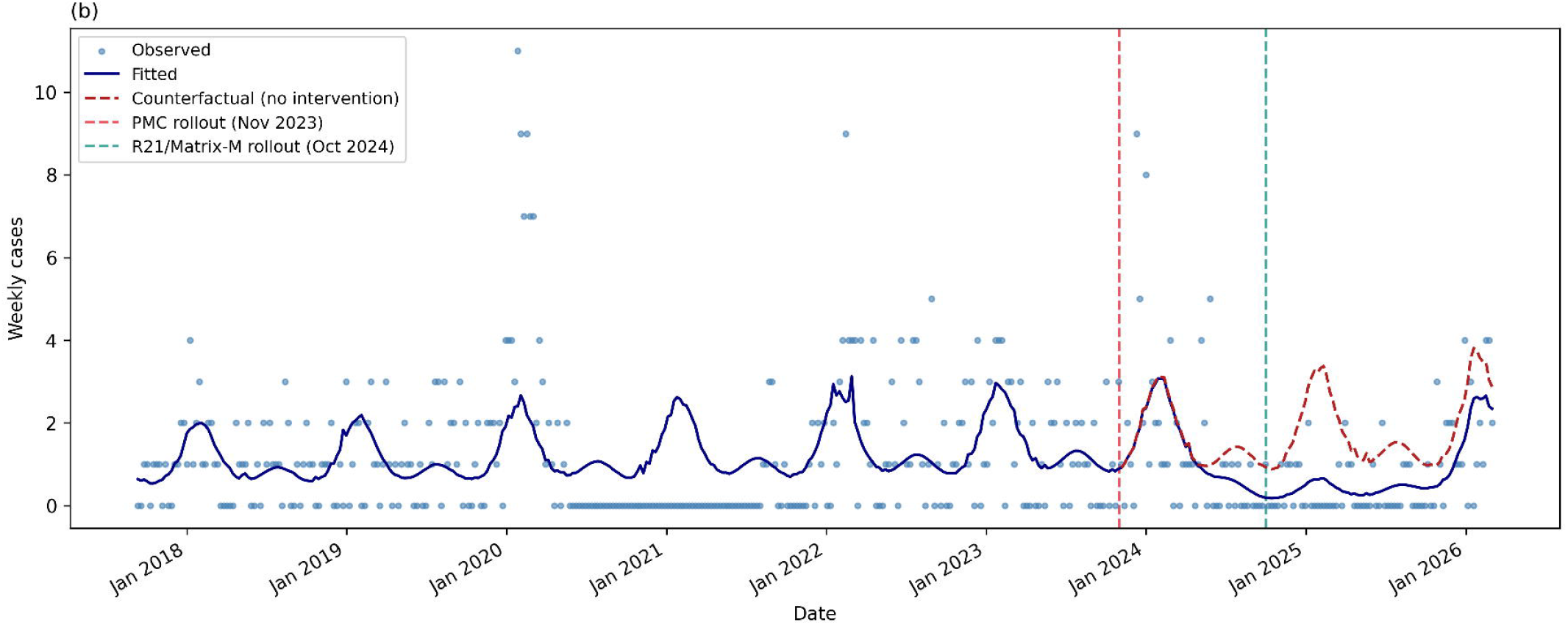
Interrupted time series analysis with 6-month linear coverage ramp-up and precipitation covariate (NASA POWER MERRA2). (a) Weekly malaria incidence: observed counts (blue dots), fitted series (dark line), and counterfactual projection (dashed red line, assuming no intervention). Red vertical dashed line = PMC rollout (November 2023); teal vertical dashed line = R21/Matrix-M rollout (October 2024). (b) Same layout for iNTS incidence. The ramp function models gradual coverage build-up over 6 months from each rollout date, with the slope change term beginning after full coverage is reached.

In the present ongoing PMC delivery, R21/Matrix-M (October 2024), targeting children <2 years, showed no detectable all-age malaria effect at 6-months after program initiation (IRR 0·860, 95% CI 0·204–3·625; p=0·837). This null finding is expected: the targeted age stratum represents approximately 6·9% of the catchment population, diluting the all-age signal substantially, and the limited vaccine uptake in initial months along with short follow-up period (6 months) may also contribute to this observation.

### Effect of Anti-Malaria Interventions on iNTS Incidence

With a 6-month ramp-up and precipitation adjustment, PMC was associated with a null iNTS level change at rollout (IRR 0·972, 95% CI 0·418–2·262; p=0·948), followed by a significant declining iNTS trend post-ramp (IRR 0·990/day, 95% CI 0·981–0·998; p=0·018). (Figure 1(b)) Seasonal adjustment was critical. A fully unadjusted segmented regression — binary step function, no Fourier harmonics, no precipitation term — yielded an apparent iNTS level increase at PMC rollout (IRR 2·766, 95% CI 1·453–5·267; p=0·002), a confounding artefact of the coincident November rainy-season transition. The artefact is removed by the Fourier harmonics together with the coverage-ramp specification; the precipitation covariate itself is a marginal refinement, since removing it alone shifted the level change only from 0·972 to 0·947 (95% CI 0·407–2·204; p=0·899). Full decomposition across all five nested specifications is in appendix table S4. The significant declining trend after full PMC coverage is consistent with a lagged malaria-mediated effect as population coverage built.

R21/Matrix-M introduction was associated with a non-significant iNTS level change (IRR 6·572, 95% CI 0·519–83·134; p=0·146) and a rising slope (IRR 1·015/day, 95% CI 1·006–1·024; p=0·001). The extremely wide confidence interval reflects sparse iNTS data in the six months following rollout; the slope rise likely reflects an underlying secular trend rather than a vaccine effect. Full sensitivity results by ramp period are in appendix table S1.

### Population-Level Malaria–NTS Interaction Strength

The SIRS model estimated a malaria–NTS susceptibility enhancement of ψ_M→N_ = 12·0 (95% bootstrap CI: 3·6–28·3) under the biologically constrained Mode 1 solution (ρ0 = 22·7%, 95% bootstrap CI 17·2– 38·5%). Profile likelihood confirmed that values of ψ_M→N_ < 5 are inconsistent with the data (Δ negative log-likelihood [NLL] > 38 relative to the profile likelihood optimum). An alternative Mode 2 solution exists (ψ_M→N_ ≈ 102, ρ_mal_ ≈ 80·8%, ΔNLL = −46·9 vs Mode 1 MAP) that, though a better calibration fit to the data, estimated improbable values for two key parameters: ψ_M→N_ ≈ 102 is biologically implausible, far exceeding the 2–5-fold range from clinical case-control studies; and ρ_0_ ≈ 80·8% implies detection of approximately 81% of all malaria infections by passive facility-based surveillance — incompatible with the likely malaria-infection detection ratio for a rural hyper-endemic DRC health zone setting. All solutions support that malaria infection substantially enhances the risk of NTS infection; the absolute fold-change cannot be determined from facility surveillance counts alone (see Limitations).

### Intervention Impact: Percent Reduction in iNTS Incidence

At 50% PMC coverage, R21/Matrix-M alone achieved 16–40% iNTS burden reduction depending on coverage; PMC alone achieved 7–20%; an investigational iNTS vaccine at 60% efficacy achieved 11–30%; and combined strategies achieved 25–60% reduction, with synergistic effects exceeding the sum of individual interventions. Hence a significant reduction in iNTS burden could be achieved, based on our estimations, with available malaria interventions and even without an iNTS-specific vaccine.

### Elimination Threshold Analysis

To achieve NTS elimination (<1 infection per million at year 5), an iNTS-specific vaccine with ≥70% efficacy at high coverage is necessary: coverage thresholds are ≥75% at 90% vaccine efficacy, ≥85% at80% efficacy, and ≥95% at 70% efficacy. Malaria control alone cannot achieve elimination given the large undetected NTS reservoir, approximately 2·0% (95% bootstrap CI 1·6–3·4%) of bacteraemia cases detected through surveillance (Supplement S5). Combined malaria and iNTS interventions show super-additive effects (Supplement S9) at sub-elimination coverage levels (appendix figure S1), supporting integrated deployment strategies.

## Discussion

The study provides the first population-level estimates of malaria’s causal contribution to invasive non-typhoidal *Salmonella* (iNTS) burden, combining quasi-experimental and mechanistic modelling approaches. Three principal findings emerge. First, an interrupted times series (ITS) analysis showed that the introduction of malaria control interventions in the Kisantu Health Zone was associated with a substantial decline in both malaria infection (IRR 0·197, 95% CI 0·096–0·401; p<0·001) and iNTS disease incidence (IRR 0·990/day, 95% CI 0·981–0·998; p=0·018). Second, a calibrated Susceptible-Infectious-Recovered-Susceptible (SIRS) model estimated that active or recent malaria infection enhanced the risk of acquiring NTS infection approximately 12-fold (Mode 1 maximum a *posteriori* [MAP] estimate; 95% bootstrap CI: 3·6–28·3). Third, combined R21/Matrix-M vaccine and perennial malaria chemoprevention (PMC) interventions were projected to reduce iNTS disease burden by 16–37%, with a range driven by assumed vaccine and PMC coverage levels (Figure 2). Full elimination of NTS transmission was only achieved in scenarios incorporating an iNTS-specific vaccine with efficacy ≥70% at high coverage.

**Figure 2.**
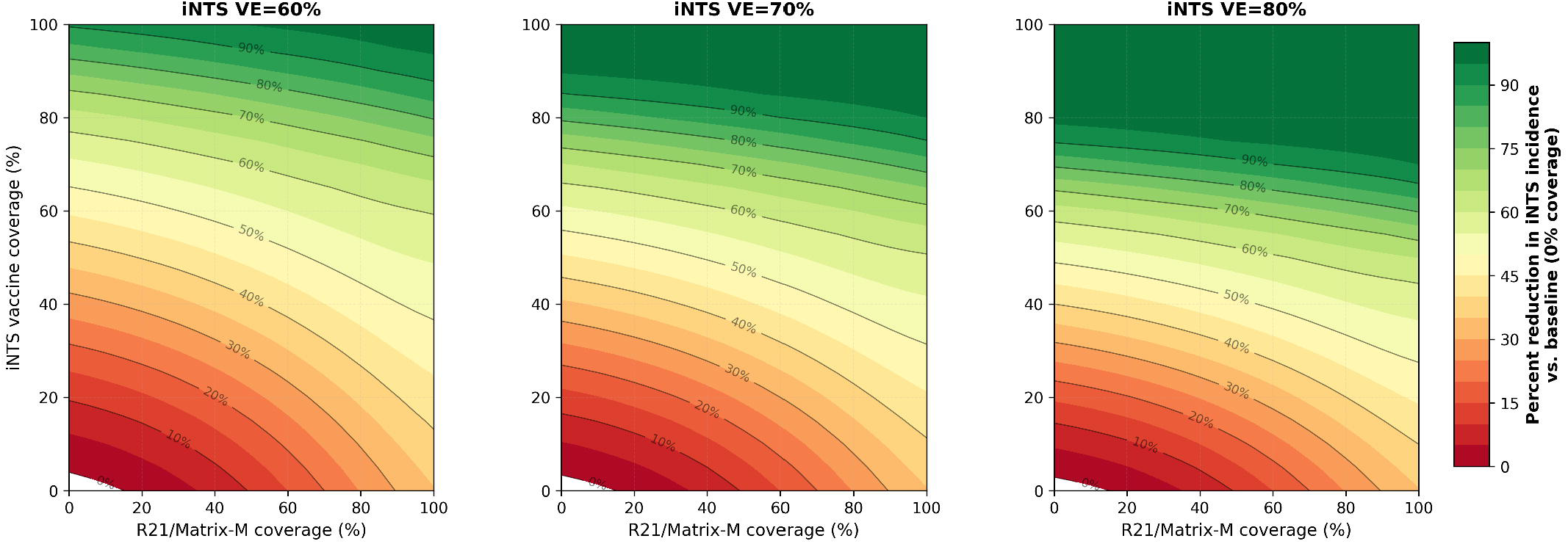
Percent reduction in iNTS incidence from baseline as a function of R21/Matrix-M coverage (x-axis) and iNTS vaccine coverage (y-axis), at PMC fixed at 50% coverage. Three panels show iNTS vaccine efficacies of 60%, 70%, and 80%. Contour lines at 10% intervals indicate combinations achieving equivalent burden reduction; darker green indicates greater reduction.

These findings quantify, at population-level, the mediating role of malaria infection in iNTS disease, previously described in a convergent body of literature.^8, 23^ Our estimated 12-fold increase in iNTS disease’s risk associated with active or recent malaria infection exceeds the 2–5-fold range estimated in a longitudinal case-control analysis looking at the incidence of all bacteremia associated with malaria parasitemia.^11^ Our estimated malaria incidence reduction following PMC introduction (IRR 0·197, 95% CI 0·096–0·401; p<0·001) is larger than the approximate 50% reduction reported pre- and post-PMC implementation in Nigeria in 2021-22.^24^ It is also larger than other published estimates of the Seasonal Malaria Chemoprevention (SMC) impact on the occurrence of malaria in West and Central Africa.^25–27^ Because underlying study designs, populations, and surveillance systems differ across settings, these estimates are not directly comparable, and differences should be interpreted qualitatively rather than as formal tests of difference. To our knowledge, no previous study has estimated the population-level impact of malaria control measures on iNTS burden.

Adjusting for seasonality was critical to these estimates. In a fully unadjusted model carrying no seasonal control, PMC introduction was apparently associated with an increased iNTS incidence (IRR 2·766, 95% CI 1·453–5·267; p=0·002); this reflected confounding by the coincident November rainy-season transition rather than a true intervention effect and was eliminated once Fourier harmonics and the coverage ramp were included (IRR 0·972, 95% CI 0·418–2·262; p=0·948). The same fully unadjusted model also produced a spurious malaria level increase at the R21 breakpoint (IRR 2·824, 95% CI 1·677–4·755; p<0·001) that no vaccine mechanism can explain, underscoring how strongly uncontrolled seasonality distorts these estimates. The precipitation covariate itself proved to be a marginal refinement: adding or removing it changed no level or slope estimate materially (appendix table S4). The ramp specification, modelling gradual six-month coverage build-up, together with the Fourier seasonal harmonics, separated the intervention signal from seasonal confounding. The results under this specification are consistent with a lagged, malaria-mediated effect on iNTS incidence. A formal causal mediation analysis, intended to quantify the proportion of this effect operating through malaria, did not provide additional support: bootstrap confidence intervals were wide at all lags tested (0-28 days) and no stable mediation signal was detected (appendix table S2, appendix figure S4b), most likely reflecting sparse iNTS case counts. We therefore base our inference of a malaria-mediated pathway primarily on the calibrated SIRS model and the ITS results rather than on the mediation analysis and treat the mediation findings as exploratory. Multi-site studies with larger case counts are needed to test mediation formally.

These findings provide evidence consistent with a synergistic benefit of malaria control interventions for both malaria and iNTS disease containment. In the absence of a licensed iNTS vaccine, our modelling suggests that a substantial proportion of iNTS disease burden (16-37%) could be reduced by combining PMC and malaria vaccination. The uptake of licensed malaria vaccines remains uneven across the region: in 2025, WHO-reported first-dose coverage of R21/Matrix-M vaccine averaged 67% (range 15% in Niger to 93% in Guinea), with above-average coverage of 86% in the Democratic Republic of Congo (DRC).^28^ Countries with high burden of both malaria and iNTS disease, such as the DRC, should prioritize the introduction of a licensed malaria vaccine into routine immunisation programmes where not yet in place, and policies to raise coverage where these programmes already exist. Targeted perennial and seasonal malaria chemoprevention in high-incidence regions could provide additional benefits on the burden of malaria and mediated infections. However, reducing host susceptibility to iNTS disease through malaria control on its own is unlikely to eliminate NTS transmission at population level, as it does not act on environmental or zoonotic transmission sources. Our modelling indicates that full elimination of NTS infection would require an iNTS-specific vaccine with ≥70% efficacy at high coverage, underscoring that malaria control and iNTS-specific interventions should be viewed as complementary rather than substitutable. Previous modeling studies considered only NTS transmission, limiting the ability to assess the extent of potential malaria prevention effects on iNTS risk.^6, 29^

Several limitations warrant careful consideration. First, a fundamental identifiability trade-off exists between the estimated susceptibility enhancement (ψ_M→N_) and surveillance reporting fractions (ρ_0_, ρ_1_): facility-based case counts alone cannot uniquely identify both parameters. The existence of two distinct solution modes (Mode 1 and Mode 2) in our calibration illustrates this trade-off. A sensitivity analysis fixing ψ_M→N_ at 3·0, consistent with prior literature (appendix S11), fitted the data substantially worse than the primary model (ΔNLL = +39·5 relative to the Mode 1 MAP), indicating that the data support ψM→N well above 3·0. Resolving this ambiguity more definitively will require external data — population-based blood culture surveys, care-seeking surveys, or healthcare utilisation data — to independently constrain ρ_0_ and ρ_1_, and our ψ_M→N_ estimate should be interpreted with this uncertainty in mind. Second, the iNTS vaccine modelled does not yet exist; the efficacy sweep (30–90%) characterises a theoretical elimination boundary under plausible future scenarios rather than predicting real-world outcomes of a specific product. Third, the model assumes a homogeneous catchment population; spatial clustering of NTS risk, plausible given known heterogeneity in water, sanitation, and hygiene access, could alter local elimination thresholds and was not captured here. Fourth, reporting fractions were estimated from surveillance data and could be confounded by temporal changes in surveillance intensity or healthcare-seeking behavior that we could not fully disentangle from true epidemiological change. Fifth, unmeasured co-infection modifiers — including nutritional status, Human Immunodeficiency Virus (HIV) infection, and sickle cell trait — may partly explain the high estimated ψ_M→N_ and were not accounted for in the model. Sixth, calibration to a single health zone limits generalizability; the DRC-specific policy recommendations above should be treated as illustrative rather than directly transferrable, and other African settings would require adapted parameterisation based on local transmission intensity and surveillance characteristics. Multi-site studies spanning settings with differing malaria and iNTS endemicity are needed to validate and generalize these findings before they inform region-wide policy. Finally, we did not conduct a cost-effectiveness analysis of the interventions modelled; incorporating local costs, health system capacity and competing health priorities indicators would strengthen the evidence base for procurement and budgeting decisions and is an important direction for future work.

## Conclusion

In summary, we document population-level surveillance evidence for a malaria–iNTS interaction using interrupted time series and mechanistic modelling. After accounting for a 6-month immunisation coverage ramp-up and precipitation-driven seasonality, PMC was associated with an 80% reduction in malaria incidence at full coverage (IRR 0·197, 95% CI 0·096–0·401; p<0·001) and a significant decline in iNTS trend (IRR 0·990/day, 95% CI 0·981–0·998; p=0·018), a pattern consistent with a lagged, malaria-mediated effect on iNTS disease. Our calibrated mechanistic model estimates an approximate 12-fold enhancement of NTS susceptibility associated with malaria infection (ψM→N = 12·0, Mode 1 MAP, 95% CI: 3·6–28·3), an estimate that should be interpreted alongside the identifiable caveats described above. These findings support the prioritisation of malaria control interventions roll out and coverage expansion, both for their direct benefits on malaria burden and for their probable co-benefits on iNTS disease burden, while highlighting that full elimination of NTS disease will additionally require a high-efficacy iNTS-specific vaccine; environmental control strategies, such as WASH; and validation of these findings across multiple settings.

## Contributors

Jonathan D. Sugimoto: Conceptualization, Methodology, Formal analysis, Writing – original draft. Camille Dauvergne: Conceptualization, Methodology, Formal analysis, Funding acquisition, Investigation, Writing – original draft. Jean−Paul Kumbukama: Investigation, Methodology, Formal analysis, Writing – original draft. Mohamadou Siribie: Funding acquisition, Investigation, Validation. Marie−France Phoba: Investigation, Supervision. Jules Mbuyamba: Investigation, Supervision. Seung Eun Kyung: Data curation, Investigation. Eunjeong Jeong: Data curation, Investigation. Wonchul Chung: Data curation, Investigation. Andreia Regina Augusto dos Santos: Investigation. Rita Soares Barbosa Cardona: Investigation. Hyonjin Jeon: Resources, Funding acquisition. Jessica Cowden: Methodology, Validation. Florian Marks: Conceptualization, Funding acquisition, Investigation, Validation, Supervision, Writing – review & editing. Octavie Lunguya: Conceptualization, Investigation, Validation, Supervision, Writing – review & editing. Birkneh T. Tadesse: Conceptualization, Funding acquisition, Investigation, Methodology, Validation, Supervision, Writing – review & editing.

## Declaration of interests

We declare no competing interests.

## Data sharing

Individual-level surveillance data are held under data-sharing agreement with the Institut National de Recherche Biomédicale (INRB), Kinshasa, DRC, and cannot be made publicly available. Deidentified aggregate weekly case counts and analysis code will be made available at https://github.com/jsugimot/drc_malaria_ints_ivi.git. Requests for access to individual-level data should be directed to the corresponding author.

## Acknowledgements

This research was funded by the Gates Foundation (INV-047715, INV-007844, INV-082265, INV-077454), the European and Developing Countries Clinical Trials Partnership (RIA2017S-2027), and the Swedish Styrelsen för internationellt utvecklingssamarbete (17106).

## Artificial intelligence declaration

Development and implementation of the analysis workflow were assisted by an agentic artificial intelligence system (Claude for Windows, version 1·1·9669, Anthropic PBC). The AI system assisted with code generation under the direct supervision and scientific direction of the first authors, who take full responsibility for the integrity of the analysis and accuracy of all reported results.

## Notes

### Competing Interest Statement

The authors have declared no competing interest.

### Author Declarations

Institutional Review Board of the International Vaccine Institute gave ethical approval for this work. Ethics Review Committee of the School of Public Health of Kinshasa, DRC, gave ethical approval for this work.

## References

1. Marks F, von Kalckreuth V, Aaby P. Incidence of invasive Salmonella disease in sub-Saharan Africa: a multicentre population-based surveillance study. Lancet Glob Health. 2017;5(3):e310–e23.

2. Feasey NA, Dougan G, Kingsley RA, Heyderman RS, Gordon MA. Invasive non-typhoidal Salmonella disease: an emerging and neglected tropical disease in Africa. Lancet. 2012;379(9835):2489–99.

3. Marchello CS, Dale AP, Pisharody S, Rubach MP, Crump JA. Prevalence of community-onset bloodstream infections in low- and middle-income countries: a systematic review and meta-analysis. Antimicrob Agents Chemother. 2020;64(4):e01974–19.

4. He Y, Jia Q, Cai K. The global, regional, and national burden of invasive non-typhoidal Salmonella (iNTS): an analysis from the Global Burden of Disease Study 1990-2021. PLoS Negl Trop Dis. 2025;19(4):e0012960.

5. Crump JA, Sjolund-Karlsson M, Gordon MA, Parry CM. Epidemiology, clinical presentation, laboratory diagnosis, antimicrobial resistance, and antimicrobial management of invasive Salmonella infections. Clin Microbiol Rev. 2015;28(4):901–37.

6. Cassese D, Dimitri N, Breghi G, Spadafina T. Effectiveness of iNTS vaccination in sub-Saharan Africa. Sci Rep. 2025;15(1):3765.

7. Tennant SM, MacLennan CA, Simon R, Martin LB, Khan MI. Nontyphoidal salmonella disease: Current status of vaccine research and development. Vaccine. 2016;34(26):2907–10.

8. Park SE, Pak GD, Aaby P, Adu-Sarkodie Y, Ali M, Aseffa A, et al. The Relationship Between Invasive Nontyphoidal Salmonella Disease, Other Bacterial Bloodstream Infections, and Malaria in Sub-Saharan Africa. Clin Infect Dis. 2016;62 Suppl 1(Suppl 1):S23–31.

9. Krumkamp R, Kreuels B, Sarpong N, Boahen KG, Foli G, Hogan B, et al. Association Between Malaria and Invasive Nontyphoidal Salmonella Infection in a Hospital Study: Accounting for Berkson’s Bias. Clin Infect Dis. 2016;62 Suppl 1:S83–9.

10. Cunnington AJ, de Souza JB, Walther M, Riley EM. Malaria impairs resistance to Salmonella through heme- and heme oxygenase-dependent dysfunctional granulocyte mobilization. Nat Med. 2012;18(1):120–7.

11. Scott JAG, Berkley JA, Mwangi I. Relation between falciparum malaria and bacteraemia in Kenyan children. Lancet. 2011;378(9799):1316–23.

12. Berkley JA, Lowe BS, Mwangi I. Bacteremia among children admitted to a rural hospital in Kenya. N Engl J Med. 2005;352(1):39–47.

13. Datoo MS, Dicko A, Tinto H. Safety and efficacy of malaria vaccine candidate R21/Matrix-M in African children: a multicentre, doubleblind, randomised, phase 3 trial. Lancet. 2024;403(10426):533–44.

14. Chen RTQ, Rubanova Y, Bettencourt J, Duvenaud D, editors. Neural ordinary differential equations. Advances in Neural Information Processing Systems (NeurIPS); 2018.

15. von Kalckreuth V, Konings F, Aaby P. The Typhoid Fever Surveillance in Africa Program (TSAP): Clinical, Diagnostic, and Epidemiological Methodologies. Clin Infect Dis. 2016;62(Suppl 1):S9–S16.

16. Park SE, Toy T, Cruz Espinoza LM. The Severe Typhoid Fever in Africa Program: Study Design and Methodology to Assess Disease Severity, Host Immunity, and Carriage Associated With Invasive Salmonellosis. Clin Infect Dis. 2019;69(Suppl 6):S422–S34.

17. Newey WK, West KD. A simple, positive semi-definite, heteroskedasticity and autocorrelation consistent covariance matrix. Econometrica. 1987;55(3):703–8.

18. Newey WK, West KD. Automatic lag selection in covariance matrix estimation. Review of Economic Studies. 1994;61(4):631–53.

19. VanderWeele TJ. Explanation in Causal Inference: Methods for Mediation and Interaction. New York, NY: Oxford University Press; 2015.

20. Kidger P. On neural differential equations [PhD thesis]: University of Oxford; 2021.

21. Cairns M, Roca-Feltrer A, Garske T. Estimating the potential public health impact of seasonal malaria chemoprevention in African children. Nat Commun. 2012;3:881.

22. Dye C. Global epidemiology of tuberculosis. Lancet. 2006;367(9514):938–40.

23. Nyirenda TS, Mandala WL, Gordon MA, Mastroeni P. Immunological bases of increased susceptibility to invasive nontyphoidal Salmonella infection in children with malaria and anaemia. Microbes Infect. 2018;20(9-10):589–98.

24. Ikechukwu EC, Okereke E, Ogunmola O, Chukwumerije J, Emeto D, Salifu E, et al. Impact of seasonal malaria chemoprevention: a plausibility evaluation of routine data from health facilities in three implementing states in Nigeria. Malar J. 2025;24(1):396.

25. Kirakoya-Samadoulougou F, De Brouwere V, Fokam AF, Ouédraogo M, Yé Y. Assessing the effect of seasonal malaria chemoprevention on malaria burden among children under 5 years in Burkina Faso. Malar J. 2022;21(1):143.

26. Effectiveness of seasonal malaria chemoprevention at scale in west and central Africa: an observational study. Lancet. 2020;396(10265):1829–40.

27. Huang S, Baker K, Ibinaiye T, Oresanya O, Nnaji C, Richardson S. Impact of seasonal malaria chemoprevention based on the number of medicines doses received on malaria burden among children aged 3-59 months in Nigeria: A propensity score-matched analysis. Trop Med Int Health. 2024;29(8):668–79.

28. World Health Organization. Malaria vaccination coverage Geneva: WHO Immunization Data portal (WIISE); 2026 [Available from: https://immunizationdata.who.int/global/wiise-detail-page/malaria-vaccination-coverage.

29. Mastroeni P, Rossi O. Immunology, epidemiology and mathematical modelling towards a better understanding of invasive non-typhoidal Salmonella disease and rational vaccination approaches. Expert Rev Vaccines. 2016;15(12):1545–55.

