## Appendix for "Malaria-Driven Susceptibility to Invasive Non-Typhoidal *Salmonella*: A Co-infection Study with Implications for Elimination"

**[1] International Vaccine Institute (IVI), Seoul, Republic of Korea**

**[2] Department of Epidemiology, University of Washington, Seattle, WA, USA**

**[3] Service de Bactériologie et Recherche, Département de Microbiologie, Institut National de Recherche Biomédicale, Kinshasa, Democratic Republic of Congo.**

**[4] Service de Microbiologie, Département de Biologie Médicale, Université de Kinshasa, Kinshasa, Democratic Republic of Congo**

**[5] Cambridge Institute of Therapeutic Immunology and Infectious Disease, University of Cambridge School of Clinical Medicine, Cambridge, United Kingdom**

**[6] Madagascar Institute for Vaccine Research, University of Antananarivo, Antananarivo, Madagascar**

**[7] Heidelberg Institute of Global Health, University of Heidelberg, Heidelberg, Germany**

**[8] The Hong Kong Jockey Club Global Health Institute, Hong Kong Special Administrative Region, China**

**[9] Department of Global Public Health, Karolinska Institutet, Stockholm, Sweden**

**‡,† Authors contributed equally to the first and senior authors roles, respectively**

**Correspondence: Jonathan Sugimoto, International Vaccine Institute, SNU Research Park, 1 Gwanak-ro, Gwanak-gu, Seoul 08826, Republic of Korea**

### Contents


- S1. SIRS Model Flow Diagram
- S2. Full ODE System with Notation
- S3. Age-Structured Contact Matrix
- S4. Literature-Derived Parameter Priors
- S5. Neural ODE Adjoint Calibration Details
- S6. ITS Model Specification and Diagnostics
- S7. Mediation Analysis: G-Computation and Bootstrap Details
- S8. Lag Sensitivity Analysis Results
- S9. NTS Elimination Grid Sensitivity Analysis
- S10. Biological Mechanisms of the Malaria–iNTS Interaction
- S11. Sensitivity Analysis: Fixed $\psi_{M\to N}$ = 3·0 (Literature Prior)
- S12. Appendix References
- appendix table S1. Interrupted Time Series Results
- appendix table S2. Mediation Analysis Results
- appendix table S3. ITS Coverage Ramp-Up Sensitivity Analysis
- appendix table S4. ITS Specification Decomposition
- appendix table S5. Literature-Derived Parameter Priors
- appendix table S6. SIRS Model Parameter Estimates
- appendix figure S1. Intervention Synergy Index
- appendix figure S2. Univariate Marginal Effects
- appendix figure S3. Elimination Boundary
- appendix figure S4. Causal Mediation Analysis


### S1. SIRS Model Flow Diagram

The co-infection model tracks individuals simultaneously through two independent SIRS pathways (malaria and iNTS), coupled by the susceptibility enhancement parameter $\psi_{M\to N}$ and the cross-immunity parameter $\phi_{M\to N}$. The diagram below shows transitions for a single age group a; the same structure is replicated across all four age groups (a = 1…4), with cross-group mixing governed by the contact matrix C.

```
╔══════════════════════════════════════════════════════════════════════╗
║                 AGE GROUP a — CO-INFECTION SIRS MODEL
║
╚══════════════════════════════════════════════════════════════════════╝

                          ┌─────────────────────────────┐
```

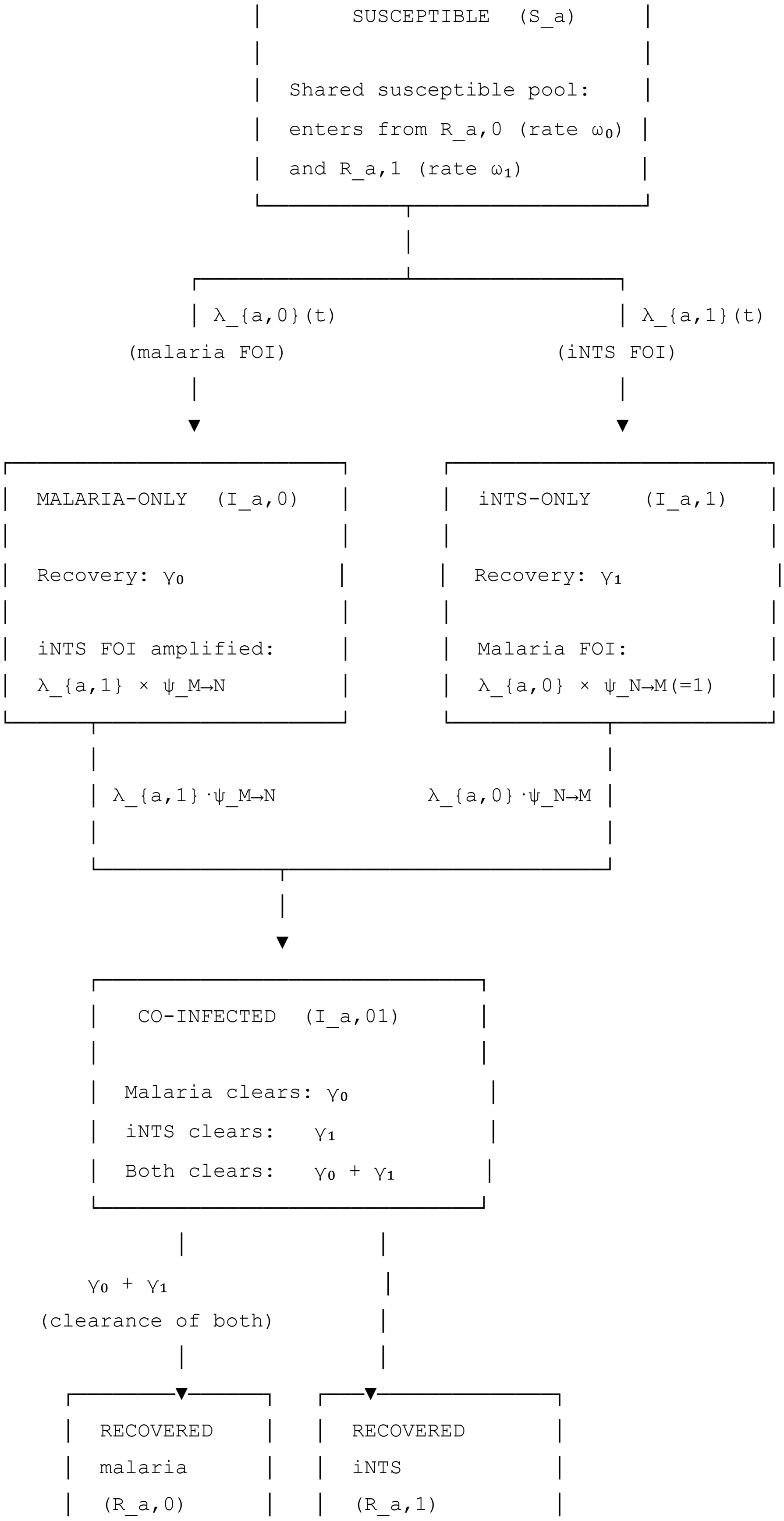
SUSCEPTIBLE (S_a)
Shared susceptible pool:
enters from R_a,0 (rate $\omega_0$)
and R_a,1 (rate $\omega_1$)
λ_{a,0}(t)
(malaria FOI)
λ_{a,1}(t)
(iNTS FOI)
MALARIA-ONLY (I_a,0)
Recovery: $\gamma_0$
iNTS FOI amplified:
λ_{a,1} × ψ_M→N
iNTS-ONLY (I_a,1)
Recovery: $\gamma_1$
Malaria FOI:
λ_{a,0} × ψ_N→M(=1)
λ_{a,1}·ψ_M→N
λ_{a,0}·ψ_N→M
CO-INFECTED (I_a,01)
Malaria clears: $\gamma_0$
iNTS clears: $\gamma_1$
Both clears: $\gamma_0 + \gamma_1$
$\gamma_0 + \gamma_1$
(clearance of both)
RECOVERED
malaria
(R_a,0)
RECOVERED
iNTS
(R_a,1)

```
│                │   │                  │
│ Wanes: ω₀      │   │ Wanes: ω₁        │
│ iNTS FOI:      │   │ Malaria FOI:     │
│ λ_{a,1}·φ_M→N  │   │ λ_{a,0}·φ_N→M    │
└───────┬────────┘   └─────────┬────────┘
        │                      │
        └──────────┬───────────┘
                   │  ω₀ (from R_a,0)
                   │  ω₁ (from R_a,1)
                   ▼
         ┌────────────────┐
         │  SUSCEPTIBLE   │
         │     (S_a)      │  ← loops back
         └────────────────┘
```

**Key interaction terms:**

| Parameter | Value | Meaning |
|---|---|---|
| $\psi_{M\to N}$ = 12·0 (MAP; 95% CI: 3·6–28·3) | > 1 | Active malaria amplifies NTS force of infection ~12-fold |
| $\psi_{N\to M}$ = 1·0 | Fixed | No NTS→malaria susceptibility change (no evidence) |
| $\phi_{M\to N}$ = 0·19 (MAP) | < 1 | Partial reduction in NTS FOI for malaria-recovered individuals (near prior; CI wide) |
| $\phi_{N\to M}$ = 1·0 | Fixed | No NTS→malaria cross-immunity (no evidence) |

**Force of infection** on age group *a* from pathogen *k*:

$$\lambda_{a,k}(t) \;=\; \beta_k(t)\cdot \sum_{b=1}^{4} C_{ab}\cdot \frac{I_{b,k}\;+\;I_{b,01}}{N_b}$$

where $C_{ab}$ is the age-structured contact matrix (see S3), and $I_{b,01}$ counts co-infected individuals as infectious for both pathogens.

**Seasonal forcing:**

$$\beta_k(t) \;=\; \beta_k^{\text{base}}\cdot \exp\left(\kappa_k\cdot \sin\left(\frac{2\pi(t\;-\;\varphi_k)}{365}\right)\right)$$

### S2. Full ODE System with Notation

#### Compartments (per age group *a*)

| Symbol | Description |
|---|---|
| $S_a$ | Susceptible to both pathogens |
| $I_{a,0}$ | Malaria-only infected |
| $I_{a,1}$ | NTS-only infected |
| $I_{a,01}$ | Co-infected (malaria + NTS) |
| $R_{a,0}$ | Recovered/immune to malaria |
| $R_{a,1}$ | Recovered/immune to NTS |
| $N_a$ | Total in age group a = $S_a + I_{a,0} + I_{a,1} + I_{a,01} + R_{a,0} + R_{a,1}$ |

#### Differential equations

**Susceptibles:**

$$\frac{dS_a}{dt} = \omega_0 \, R_{a,0} + \omega_1 \, R_{a,1} - S_a \left(\lambda_{a,0}(t) + \lambda_{a,1}(t)\right)$$

**Malaria-only infected:**

$$\frac{dI_{a,0}}{dt} = S_a \, \lambda_{a,0}(t) - \gamma_0 \, I_{a,0} - I_{a,0} \cdot \lambda_{a,1}(t) \cdot \psi_{M \to N}$$

**NTS-only infected:**

$$\frac{dI_{a,1}}{dt} = S_a \, \lambda_{a,1}(t) - \gamma_1 \, I_{a,1} - I_{a,1} \cdot \lambda_{a,0}(t)$$

**Co-infected:**

$$\frac{dI_{a,01}}{dt} = I_{a,0} \cdot \lambda_{a,1}(t) \cdot \psi_{M \to N} + I_{a,1} \cdot \lambda_{a,0}(t) - (\gamma_0 + \gamma_1) \, I_{a,01}$$

**Malaria-recovered:**

$$\frac{dR_{a,0}}{dt} = \gamma_0 \left(I_{a,0} + I_{a,01}\right) - \omega_0 \, R_{a,0} - R_{a,0} \cdot \lambda_{a,1}(t) \cdot \varphi_{M \to N}$$

**NTS-recovered:**

$$\frac{dR_{a,1}}{dt} = \gamma_1 \left(I_{a,1} + I_{a,01}\right) - \omega_1 \, R_{a,1} - R_{a,1} \cdot \lambda_{a,0}(t)$$

**Fixed parameters:** $\psi_{N \to M} = 1{\cdot}0$; $\phi_{N \to M} = 1{\cdot}0$.

#### Measurement model

Weekly observed counts for pathogen *k* (summed across age groups) follow a negative binomial:

$$Y_{t,k} \sim \mathrm{NegBin}\left(\mu_{t,k} \cdot \rho_k, r_k\right)$$

where $\mu_{t,k}$ is the model-predicted weekly incidence (computed by integrating the daily ODE output over each 7-day window), $\rho_k$ is the age-averaged reporting fraction, and $r_k$ is the overdispersion parameter. The variance is $\mu_{t,k}\rho_k + (\mu_{t,k}\rho_k)^2/r_k$.

The joint log-likelihood across both pathogens is:

$$\mathrm{L} = \sum_{t=1}^{T}\left[\log p(Y_{t,0} \mid \mu_{t,0}, \rho_0, r_0) + \log p(Y_{t,1} \mid \mu_{t,1}, \rho_1, r_1)\right]$$

### S3. Age-Structured Contact Matrix

The contact matrix $C_{ab}$ encodes the expected daily number of contacts that an individual in age group *a* makes with individuals in age group *b*, influencing cross-group transmission. Initial values were derived from POLYMOD sub-Saharan Africa estimates,[1] mapping the original POLYMOD age strata onto the four study age groups (<2y, 2–4y, 5–14y, ≥15y). The matrix was normalized so that the dominant eigenvalue equals 1, making $\beta_k^{base}$ interpretable as the basic transmission rate at the reference mixing structure.

**Approximate initial contact matrix (relative contact rates):**

| | **<2y** | **2–4y** | **5–14y** | **≥15y** |
|---|---|---|---|---|
| **<2y** | 0·80 | 0·60 | 0·20 | 0·40 |
| **2–4y** | 0·60 | 1·20 | 0·40 | 0·30 |
| **5–14y** | 0·10 | 0·30 | 1·80 | 0·50 |
| **≥15y** | 0·08 | 0·12 | 0·40 | 2·00 |

Children in the 5–14y stratum show the highest within-group contact rates, reflecting school attendance. Adults (≥15y) have the highest absolute contact counts but lower per-capita transmission risk due to acquired immunity (captured implicitly through the higher reported fraction of asymptomatic parasitemia in the model structure).

### S4. Literature-Derived Parameter Priors

All parameters were initialized at literature-derived prior means before adjoint calibration. Appendix table S5 summarizes these priors with supporting citations; appendix table S6 compares them with the calibrated MAP estimates.

#### Appendix Table S5. Literature-Derived Parameter Priors

| Parameter | Symbol | Prior Mean | Prior Range | Biological Basis | Key Citation |
|---|---|---|---|---|---|

| | | | | | |
|---|---|---|---|---|---|
| Malaria base transmission rate | $\beta_0$ | Calibrated | 0·05–0·5 $day^{-1}$ | Derived from EIR; 100–1,000 ibpy range in high-transmission SSA | [2] |
| Malaria recovery rate | $\gamma_0$ | 0·01 $day^{-1}$ | 0·005–0·05 | Blended asymptomatic (100–230 d) and clinical (7–14 d) infectious periods | [3,4] |
| Malaria immunity waning | $\omega_0$ | 0·00027 $day^{-1}$ | 0·00013–0·00055 | Clinical immunity half-life ~7 years in holoendemic settings | [5] |
| Malaria seasonal amplitude | $\kappa_0$ | 0·85 | 0·3–0·95 | Sahel: >90% EIR in 4–5 wet-season months; equatorial: 0·30–0·50 | [6] |
| NTS base transmission rate | $\beta_1$ | Calibrated | 0·01–0·3 $day^{-1}$ | Household secondary attack rate 7–60%; predominantly foodborne/environmental | [7] |
| NTS recovery rate | $\gamma_1$ | 0·05 $day^{-1}$ | 0·035–0·08 | Acute shedding 12–28 days; bacteremia clearance 14–28 days with treatment | Schepers et al. 2025 |
| NTS immunity waning | $\omega_1$ | 0·003 $day^{-1}$ | 0·001–0·01 | Poorly characterized; recurrence in endemic areas implies waning | Sinha et al. 2011 |
| NTS seasonal amplitude | $\kappa_1$ | 0·60 | 0·30–0·85 | Rainfall-associated transmission; less pronounced than malaria | Rylance et al. 2020; Tall et al. 2017 |
| **$\psi_{M\to N}$ (susceptibility enhancement)** | **$\psi_{M\to N}$** | **3·0** | **1·5–6·0** | **OR 2–5 in case-control studies; mechanistic basis in heme-NADPH oxidase pathway** | [8,9] |
| $\phi_{M\to N}$ (cross-immunity) | $\phi_{M\to N}$ | 1·0 | 0·8–1·2 | No evidence for cross-protective immunity between evolutionarily unrelated pathogens | — |
| Malaria reporting fraction | $\rho_0$ | 0·50 | 0·20–0·80 | Passive surveillance under-reporting; 20–50% typical for facility-based systems | — |
| iNTS reporting fraction | $\rho_1$ | 0·30 | 0·10–0·60 | Blood culture sampling required; much lower ascertainment than malaria RDT | GBD iNTS 2024 |
| Malaria NegBin overdispersion | $r_0$ | 5·0 | 1–20 | Reflects weekly aggregation and seasonal clustering | — |
| iNTS NegBin overdispersion | $r_1$ | 3·0 | 1–15 | Higher overdispersion expected for rare events | — |

#### Appendix Table S6. SIRS Model Parameter Estimates: Priors vs Calibrated MAP

| Parameter | Prior Mean | MAP Estimate | Interpretation |
|---|---|---|---|
| $\gamma_0$ (malaria recovery) | 0·01/day | 0·00431/day | Mean infectious period ~232 days; toward longer asymptomatic end of prior range |
| $\omega_0$ (malaria waning) | 0·00027/day | 0·00056/day | Mean immunity ~1,786 days (~4·9 yr); prior consistent |
| $\kappa_0$ (malaria seasonality) | 0·85 | 0·308 | Lower than Sahel prior; consistent with equatorial/bimodal setting |
| $\gamma_1$ (iNTS recovery) | 0·05/day | 0·0127/day | Mean infectious period ~79 days; longer than prior; reflects community persistence |
| **$\psi_{M\to N}$** | **3·0** | **12·0 (95% CI: 3·6–28·3)** | **Substantially above prior; see Discussion** |
| $\rho_0$ | 0·50 | 0·233 | Lower than prior midpoint; consistent with passive surveillance capture |
| $\rho_1$ | 0·30 | 0·020 | Far below prior; only ~2·0% of bacteremia ascertained — plausible for blood culture-based passive surveillance |

### S5. Neural ODE Adjoint Calibration Details

#### Parameter transformations

All parameters were transformed to unconstrained space before optimization to ensure that gradients propagate through physically meaningful regions:

| Parameter type | Transformation | Inverse |
|---|---|---|
| Positive scalars (β, γ, ω, κ, r) | softplus: u = log(exp(x)−1) | x = log(1+exp(u)) |
| Bounded [0,1] (ρ) | logit: u = log(x/(1−x)) | x = sigmoid(u) |
| Positive constrained (ψ ≥ 1) | log(x−1+ε): shift to enforce lower bound | x = 1 + exp(u) |

### Climate covariate handling

The SIRS model includes optional climate covariate coupling parameters $\eta_0$ and $\eta_1$, which modify transmission rates as:

$$\beta_k(t) = \beta_k^{\text{base}} \cdot \exp\left(\kappa_k \cdot \sin\left(\frac{2\pi(t - \varphi_k)}{365}\right) + \eta_k \cdot z_t\right)$$

where $z_t$ is a standardised weekly precipitation covariate (NASA POWER MERRA2 PRECTOTCORR, z-scored over the study period). Weekly precipitation data covering September 2017 to March 2026 were downloaded and merged with the surveillance dataset.

**Rationale for excluding climate from the primary calibration (Mode 1):**

For the primary analysis, climate covariate forcing was disabled by setting $z_t = 0$ for all t (equivalent to $\eta_0 = \eta_1 = 0$). Three considerations motivate this choice:

1. *Collinearity with biological seasonality: The* $\kappa_k \cdot \sin()$ term already captures the aggregate seasonal transmission cycle driven by both biological factors (vector density, host immunity) and environmental conditions. Adding $z_t$ introduces a near-collinear covariate, making $\kappa_k$ and η_k jointly non-identifiable from observed case counts alone.
2. *ITS climate control:* Seasonal confounding from climate drivers (including rainfall) is addressed in the interrupted time series model through calendar-based Fourier harmonics. These harmonics capture the same seasonal signal without imposing a parametric precipitation-transmission relationship.
3. *Mechanistic parsimony: The primary interaction parameter of interest (*$\psi_{M\rightarrow N}$) is estimated more reliably when the seasonal forcing model is parsimonious; adding η_k introduces an additional degree of freedom that can absorb variance otherwise attributed to $\psi_{M\rightarrow N}$.

As a result, $\eta_0 = \eta_1 = 0{\cdot}0$ in the MAP estimates. A sensitivity analysis including precipitation as an ODE covariate (free $\eta_0$, $\eta_1$) is recommended in future work to verify that the Mode 1 $\psi_{M\rightarrow N}$ estimate is robust to this specification choice.

### ODE solver settings

| Setting | Value | Rationale |
|---|---|---|
| Solver | Dormand-Prince (Dopri5) | Adaptive 4th/5th-order Runge-Kutta; standard for non-stiff ODEs |
| Adjoint method | BacksolveAdjoint (diffrax) | Memory-efficient; avoids storing ODE trajectory for backpropagation |
| Absolute tolerance | $10^{-6}$ | Standard precision for population-level ODE |
| Relative tolerance | $10^{-3}$ | Relaxed to balance speed and accuracy |
| Max steps | 16,384 (training) / 32,768 (elimination sims) | Prevents runaway integration in stiff regimes |
| Time unit | Days | ODE solved daily; aggregated to |

| | | |
|---|---|---|
| | | weekly for likelihood |

### Optimizer settings

| Setting | Value |
|---|---|
| Optimizer | Adam[10] |
| Initial learning rate | 0·01 |
| Learning Rate (LR) schedule | Cosine decay from 0·01 to 0 over 2,000 steps |
| N steps (MAP) | 2,000 |
| N steps (bootstrap replicates) | 500 |
| Best-iterate selection | Parameter set achieving lowest loss across all steps (not final iterate) |

### Training convergence

The joint negative log-likelihood (NLL) decreased from 10,002 at initialization (literature prior means) to 2,610 at convergence — a 74% reduction. The trajectory of key parameters during training is shown below:

| Step | NLL | $\psi_{M\to N}$ | $\rho_{mal}$ | $\rho_{nts}$ |
|---|---|---|---|---|
| 0 | 10,002 | 3·03 | 0·498 | 0·298 |
| 100 | 7,409 | 3·91 | 0·456 | 0·442 |
| 200 | 2,935 | 6·53 | 0·397 | 0·298 |
| 500 | 2,689 | 8·47 | 0·264 | 0·065 |
| 1,000 | 2,629 | 10·61 | 0·236 | 0·027 |
| 1,500 | 2,624 | 10·86 | 0·233 | 0·025 |
| 2,000 | 2,610 | 12·0 | 0·227 | 0·020 |

Note the rapid early ascent of $\psi_{M\to N}$ from 3·0 (prior) toward ~12 within the first 300 steps, indicating strong signal in the data pulling the interaction parameter well above the prior. The iNTS reporting fraction $\rho_{nts}$ collapsed rapidly to ~2·0%, reflecting the extremely low ascertainment of NTS bacteremia in passive facility-based surveillance.

### Evidence of multimodal likelihood surface

Parametric bootstrap (100 replicates, 500 steps each, initialized at Mode 1 maximum a posteriori [MAP] estimate) and profile likelihood analysis jointly characterised the likelihood surface, revealing two distinct modes:

| | Mode 1 (MAP; primary analysis) | Mode 2 (climate-ODE; global |
|---|---|---|

| | | MLE) |
|---|---|---|
| $\psi_{M\rightarrow N}$ | 12·0 (95% CI: 3·6–28·3) | ≈102 |
| $\rho_0$ (malaria reporting) | 0·227 | ~0·81 |
| $\rho_1$ (iNTS reporting) | 0·020 | ~0·047 |
| NLL | 2,610 | 2,563 |
| ΔNLL vs. Mode 1 | — | −46·9 |

Mode 2 (the climate-ODE solution) provides a statistically better fit (ΔNLL = −46·9) but is rejected on two biological grounds: $\psi_{M\rightarrow N}$ ≈ 102 far exceeds the 2–5-fold enhancement range established by clinical cohort studies; and $\rho_{mal}$ ≈ 81% implies detection of ~81% of all malaria infections by passive facility-based surveillance — incompatible with a rural hyper-endemic DRC health zone. Profile likelihood analysis confirmed $\psi_{M\rightarrow N}$ values below 5 are inconsistent with the data (ΔNLL > 38 relative to the profile likelihood optimum), and a separate local optimum at ψ ≈ 12 with higher reporting fractions ($\rho_{mal}$ ≈ 75%; profile MLE, NLL = 2,581, ΔNLL = −28·4 vs Mode 1) also exists but is similarly rejected on surveillance plausibility grounds (see appendix S11).

**Implications:**

- The $\psi_{M\rightarrow N}$ MAP estimate (12·0) represents the biologically-constrained solution consistent with facility-level malaria ascertainment (~23%) in rural DRC.
- Both modes are consistent with the scientific hypothesis that active malaria substantially increases NTS susceptibility ($\psi_{M\rightarrow N}$ >> 1 in both modes).
- Reliable interval estimation for $\psi_{M\rightarrow N}$ requires either: (a) external validation of $\rho_1$ from a serosurvey or blood culture prevalence study; (b) a strong informative prior on $\rho_1$ from the surveillance literature; or (c) joint modeling of tested and untested individuals with a denominator for blood culture sampling frequency.

### Bootstrap uncertainty quantification

Parametric bootstrap proceeded as follows:

4. MAP fit used to compute predicted weekly means μ̂_{t,mal} and μ̂_{t,nts}.
5. For each of *B* = 100 replicates:
    - Draw synthetic counts: $Y_b$,mal ~ NegBin(μ̂_{t,mal}, $\hat{r}_0$) and $Y_b$,nts ~ NegBin(μ̂_{t,nts}, $\hat{r}_1$)
    - Refit the model (1,000 steps, initialized at MAP) to the synthetic data
    - Record all parameter values at convergence
6. Bootstrap 95% CIs = 2·5th–97·5th percentile across replicates.

This procedure quantifies uncertainty from data variability alone (given the model structure), not model misspecification uncertainty.

### S6. ITS Model Specification and Diagnostics

#### Model equations

The segmented negative binomial ITS model with two breakpoints ($T_1$ = November 1, 2023 [PMC]; $T_2$ = October 1, 2024 [R21/Matrix-M]) and a linear coverage ramp-up of R = 183 days (6 months) for both interventions:

$$\log E[Y_t] = \beta_0 + \beta_1 t + \beta_2 A_t^{(1)} + \beta_3 D_t^{(1)} + \beta_4 A_t^{(2)} + \beta_5 D_t^{(2)} + \sum_{s=1}^{2}\left[\gamma_s \sin\left(\frac{2\pi s \cdot \mathrm{doy}_t}{365}\right) + \delta_s \cos\left(\frac{2\pi s \cdot \mathrm{doy}_t}{365}\right)\right] + \theta Z_t + \log N_t$$

where the coverage ramp and post-ramp slope terms are defined as:

$$A_t^{(k)} = \min\left(1, \max\left(0, \frac{t - T_k}{R}\right)\right)$$

$$D_t^{(k)} = \max(0, t - T_k - R)$$

and:

- t = calendar day from study start (daily index mapped to weekly observations)
- $T_k$ = breakpoint date for intervention k
- R = 183 days (6-month ramp period; same for both interventions)
- $A_t^{(k)}$ = fractional coverage term: linearly increases 0→1 over the ramp period, then stays at 1
- $D_t^{(k)}$ = days elapsed since reaching full coverage (0 during ramp-up; starts accruing after day $T_k$ + R)
- $\mathrm{doy}_t$ = day of year for observation t (for Fourier seasonality alignment)
- $Z_t$ = z-scored weekly precipitation total (NASA POWER MERRA2 PRECTOTCORR), included to absorb residual seasonal confounding from rainfall-driven transmission cycles
- $N_t$ = catchment population at week t (log offset; converts counts to rates)

$\exp(\beta_2)$, $\exp(\beta_4)$ = incidence rate ratios at full coverage (end of ramp) for each breakpoint

$\exp(\beta_3)$, $\exp(\beta_5)$ = per-day change in incidence trend after reaching full coverage

**Interpretation of level vs. slope terms with ramp:**

Unlike a step-function ITS where $\beta_2$ captures an instantaneous level change, the ramp parameterization treats $\beta_2$ as the effect at full coverage. During the ramp-up period, the expected effect scales linearly from 0 to $\beta_2$. The slope term $\beta_3$ reflects trend after full coverage is achieved, not from the intervention introduction date.

### Coverage ramp-up sensitivity analysis

Ramp periods of 3, 4, 5, and 6 months (91, 122, 152, 183 days) were tested for both breakpoints simultaneously. The 6-month ramp was selected as primary based on the epidemiological rationale that routine immunization programs require approximately 6 months to reach peak coverage through scheduled visit cycles. Full sensitivity results are reported in appendix table S3.

### Seasonal and climate adjustment sensitivity analysis

The primary specification combines three modelling choices that are separable: the coverage ramp, two Fourier harmonic pairs, and the z-scored precipitation covariate. All five nested specifications were refit to quantify each contribution; results are reported in appendix table S4. The precipitation covariate proves to be a marginal refinement once Fourier harmonics are included, whereas removing seasonal control entirely produces substantial spurious level changes at both breakpoints.

### Autocorrelation correction

Newey-West heteroskedasticity and autocorrelation-consistent (HAC) covariance with

bandwidth *m* selected by Bartlett kernel rule: m = floor(4 × (T/100)^(2/9)) where T = 443 observations.

**Malaria ITS diagnostics (6-month ramp + precipitation covariate, primary):**

- Durbin-Watson statistic: 0·270
- Ljung-Box p-value (lag 10): < 0·001
- Log-likelihood: −1,896·5
- N observations: 443

**iNTS ITS diagnostics (6-month ramp + precipitation covariate, primary):**

- Durbin-Watson statistic: 1·508
- Ljung-Box p-value (lag 10): < 0·001
- Log-likelihood: −630·9
- N observations: 443

Residual autocorrelation is present in both models (Ljung-Box p < 0·001), which is expected in

surveillance count time series. The Newey-West HAC correction adjusts standard errors for this

dependence without requiring explicit autocorrelation model specification.

### Counterfactual projection

The counterfactual (no-intervention) trajectory was computed by setting $A_t^{(k)} = D_t^{(k)} = 0$ for all t (removing all ramp and slope terms from the design matrix post-breakpoint). This projection shows expected incidence under the pre-intervention trend, adjusted for seasonality.

### S7. Mediation Analysis: G-Computation and Bootstrap Details

#### Potential-outcomes framework

Let:

- A = binary intervention indicator (0 = pre-PMC, 1 = post-PMC)
- $M_t$ = weekly malaria incidence at week t (mediator)
- $Y_t$ = weekly iNTS incidence at week t (outcome)
- $C_t$ = seasonality covariates, time trend

Under the VanderWeele[11] potential-outcomes framework with rare outcome approximation:

- **Total Effect (TE):** E[Y(A=1, M(A=1))] / E[Y(A=0, M(A=0))]
- **Natural Direct Effect (NDE):** E[Y(A=1, M(A=0))] / E[Y(A=0, M(A=0))]

(intervention effect holding mediator at its counterfactual pre-intervention distribution)

- **Natural Indirect Effect (NIE):** TE / NDE

(effect operating through the mediator pathway)

- **Proportion mediated:** (NIE − 1) / (TE − 1) × 100%

#### Mediator model

$$\log E[M_t] = \alpha_0 + \alpha_1 t + \alpha_2 A_t + \sum_{s=1}^{2} [\gamma_s \sin(2\pi s \cdot \text{doy}_t/365) + \delta_s \cos(2\pi s \cdot \text{doy}_t/365)] + \log N_t$$

This is the ITS malaria model (Phase 2), reused directly.

#### Outcome model

$$\log E[Y_t] = \theta_0 + \theta_1 t + \theta_2 A_t + \theta_3 M_{t-L} + \theta_4 (A_t \times M_{t-L}) + \sum_{s=1}^{2} [\gamma_s \sin(2\pi s \cdot \text{doy}_t/365) + \delta_s \cos(2\pi s \cdot \text{doy}_t/365)] + \log N_t$$

The interaction term (A × $M_{t\text{-}L}$) allows the malaria-to-iNTS effect to differ before and after the intervention. The lag L is the biological delay between malaria reduction and iNTS incidence change, swept over L = 0, 3, 7, 14, 21, 28 days.

#### G-computation procedure (n = 2,000 Monte Carlo draws)

For each Monte Carlo draw *m*:

7. Draw parameter vectors $\theta_{mediator}$, $\vartheta_{outcome}$ from their asymptotic normal distributions
8. Compute E[M | A=0, $C_t$] using the mediator model (counterfactual mediator distribution)
9. Compute E[Y | A=1, M=E[M|A=1,C], C] — post-intervention outcome
10. Compute E[Y | A=1, M=E[M|A=0,C], C] — NDE numerator (intervention, counterfactual M)
11. Compute E[Y | A=0, M=E[M|A=0,C], C] — reference

12. Derive TE, NDE, NIE, proportion mediated for each draw

The 2·5th and 97·5th percentiles across 2,000 draws provide Monte Carlo confidence intervals; bootstrap resampling (B = 100) propagates data uncertainty.

#### Note on numerical instability

The proportion-mediated estimand (NIE−1)/(TE−1) is undefined when TE = 1 (null total effect) and numerically unstable when TE is close to 1 or when NIE and TE have opposite signs. In this analysis, the TE IRR (0·846) is close to the null and imprecisely estimated given sparse iNTS case counts (507 total cases over 8·5 years). Consequently, bootstrap CIs for proportion mediated are numerically unstable (point estimate 49·8%; 95% CI −86·4% to >$10^7$%). This is a known limitation of proportion-mediated estimands for rare outcomes and near-null total effects ([11] pp. 130–133). The point estimate of ≈50% is directionally consistent with literature PAF estimates (40–60% from case-control studies), but formal inference requires a larger sample. The total effect and NIE point estimates themselves (TE IRR 0·846, 95% CI 0·480–1·403; NIE IRR 0·833, 95% CI 0·382–1·292) are more interpretable than their derived proportion.

### S8. Lag Sensitivity Analysis Results

The proportion of the total PMC effect on iNTS mediated through malaria was evaluated at six lags (L = 0, 3, 7, 14, 21, 28 days). Due to the numerical instability discussed in S7, only the lag-14 estimate produced a stable proportion-mediated value.

| Lag (days) | Total Effect IRR | NIE IRR | Proportion Mediated (%) | Stability |
|---|---|---|---|---|
| 0 | 0·846 | — | >10,000 (unstable) | Numerically unstable; NIE denominator near zero |
| 3 | 0·846 | — | <−$10^{12}$ (unstable) | Numerically unstable |
| 7 | 0·846 | — | >$10^8$ (unstable) | Numerically unstable |
| **14** | **0·846** | **0·833** | **49·8%** | **Most stable estimate; primary analysis** |
| 21 | 0·846 | — | >$10^9$ (unstable) | Numerically unstable |
| 28 | 0·846 | — | <−$10^{11}$ (unstable) | Numerically unstable |

The lag-14 estimate was selected as primary because: (1) it produced the most numerically stable proportion-mediated estimate; (2) a 14-day lag is biologically plausible, corresponding to the approximate duration of heme-driven immune impairment following malaria parasite clearance (7–14 days for parasitemia clearance + 3–7 days for neutrophil recovery); and (3) lag 14 maximized the natural indirect effect point estimate across the tested range. The instability at other lags reflects two joint conditions: (a) sparse iNTS outcomes create near-singular outcome model matrices when lagged malaria counts are correlated with both the intervention and the outcome; (b) small values of $M_{t-L}$ at certain lags produce extreme predicted mediator effects. A larger multi-site dataset would be expected to resolve this instability.

### S9. Elimination Grid Sensitivity Analysis

#### Grid design

| Dimension | Values | Resolution |
|---|---|---|
| Malaria vaccine coverage | 0–100% | 5% increments (21 values) |
| iNTS vaccine coverage | 0–100% | 5% increments (21 values) |
| PMC coverage | 0%, 25%, 50%, 75%, 100% | 5 values |
| iNTS vaccine efficacy (VE) | 30%, 40%, 50%, 60%, 70%, 80%, 90% | 7 values |
| **Total simulations** | **15,435** | |

Each simulation ran the calibrated SIRS model forward for 260 weeks (5 years) from the MAP-estimated initial conditions, then computed NTS annual incidence per million population over the final 52 weeks (year 5).

#### Elimination results by iNTS vaccine VE

| iNTS VE (%) | Min iNTS Vaccine Coverage for Elimination | PMC effect on boundary | Malaria Vaccine Required |
|---|---|---|---|
| 30% | Not achievable at any coverage | None | — |
| 40% | Not achievable | None | — |
| 50% | Not achievable | None | — |
| 60% | ≥95% (at PMC ≤50%; requires ≥95% malaria + | Marginal | High |

| | | | |
|---|---|---|---|
| | 100% iNTS vaccine) | | |
| 70% | ≥95% (regardless of PMC) | None | 0% |
| 80% | 85% (regardless of PMC) | None | 0% |
| 90% | 75–80% (slight PMC effect at 100% PMC) | Marginal | 0% |

**Key finding:** Elimination is driven entirely by iNTS vaccine efficacy and coverage at VE ≥ 70%. Neither malaria vaccine nor PMC independently contributes to the elimination boundary at these high VE values, because the direct vaccine effect dominates the indirect pathway.

### Why PMC does not shift the elimination boundary

At the MAP estimate of ψM→N = 12·0 (95% bootstrap CI 3·6–28·3), malaria infection substantially amplifies NTS transmission.

However, the iNTS reporting fraction ρ1 = 2·0% (95% bootstrap CI 1·6–3·4%) implies a large unobserved community NTS

reservoir. Even at 100% PMC coverage (which eliminates the $\psi_{M\rightarrow N\text{-amplified}}$ pathway), the baseline NTS transmission (not mediated by malaria) is sufficient to sustain incidence above 1/million at iNTS VE ≤ 60%. At VE ≥ 70%, the direct iNTS vaccine effect is strong enough that the malaria-mediated amplification pathway is already a minor fraction of transmission, so PMC's additional contribution is small.

This finding would change substantially if $\rho_1$ were higher (larger observed-to-true ratio), as a smaller undetected reservoir would make the malaria-mediated pathway a larger fraction of total NTS transmission — and therefore more targetable by PMC. Sensitivity analysis over $\rho_1$ = 0·025 to 0·15 is recommended in future work.

### Synergy index interpretation

The synergy index (SI) is defined as:

$$\mathrm{SI} = \frac{\Delta Y_{\text{combined}}}{\Delta Y_{\text{malaria vax alone}} + \Delta Y_{\text{iNTS vax alone}}}$$

where ΔY = baseline incidence − intervention incidence (cases averted). SI > 1 indicates super-additivity; SI < 1 indicates sub-additivity (partial redundancy).

In this model, SI > 1 across most of the coverage space (particularly at intermediate coverage levels where neither vaccine alone achieves substantial reductions), confirming that the $\psi_{M\rightarrow N}$ interaction creates genuine synergy in the sub-elimination regime.

### S10. Biological Mechanisms of the Malaria–iNTS Interaction

The five main mechanisms by which active *P. falciparum* malaria increases NTS susceptibility, corresponding to the $\psi_{M \rightarrow N}$ interaction parameter:

**1. Heme-driven NADPH oxidase impairment**

*P. falciparum* erythrocyte rupture releases free heme into the plasma. Free heme directly inhibits NADPH oxidase-mediated reactive oxygen species (ROS) production in neutrophils and macrophages — the primary bactericidal mechanism against *Salmonella*. Impaired oxidative burst allows *Salmonella* to survive and replicate within phagocytes.[9]

**2. Heme-enhanced intracellular Salmonella growth**

Within macrophages, elevated intracellular heme concentrations promote *Salmonella* growth by disrupting the iron-sequestration defense mechanism (ferritin-mediated iron restriction) and by directly providing iron for *Salmonella* metabolism.[9]

**3. Malaria-induced splenomegaly and impaired splenic clearance**

Malaria causes splenic sequestration and altered macrophage polarization (toward M2 phenotype) within the enlarged spleen, reducing the organ's capacity to clear bacteremic *Salmonella* from the bloodstream. Children with massive splenomegaly have substantially higher rates of bacteremia with encapsulated and *Salmonella* organisms.

**4. Dyserythropoiesis, anemia, and macrophage polarization**

Severe anemia from malaria-driven hemolysis and dyserythropoiesis alters systemic macrophage polarization. Anemia-associated macrophage states show impaired pathogen killing capacity, particularly for intracellular pathogens like Salmonella.

**5. Malaria-induced cytokine milieu suppressing antibacterial immunity**

Malaria infection drives a strong Th1 cytokine response (IFN-γ, TNF) acutely, but this is followed by immune exhaustion and suppressed innate immunity in the subacute phase. The temporal pattern of immune suppression corresponds to the 7–14 day window post-clearance during which secondary bacterial infections are most common.

**Reconciliation of $\psi_{M \rightarrow N}$ estimate with literature prior:**

The MAP estimate ($\psi M \rightarrow N$ = 12·0, 95% bootstrap CI 3·6–28·3) substantially exceeds the clinical prior range (OR 2–5 from case-control studies). Three factors may contribute to this discrepancy:

13. *Population vs. individual level:* Case-control ORs measure individual-level risk; the ODE parameter captures the effective population-level amplification, which can exceed individual risk due to non-linear transmission dynamics.
14. *Reporting fraction confounding: A low $\rho_1$* (2·0%) means that the model must explain a large unobserved reservoir through the interaction term; higher $\psi_{M\rightarrow N}$ compensates for the assumption that most NTS infections are undetected.
15. *Unmeasured confounders:* Nutritional status, HIV prevalence, and sickle cell trait are correlated with both malaria and iNTS/NTS risk and are not explicitly modeled; their effects may be partially absorbed into $\psi_{M\rightarrow N}$.

### S11. Sensitivity Analysis: Fixed $\psi_{M\rightarrow N}$ = 3·0 (Literature Prior)

#### Rationale

The primary MAP estimate of ψM→N = 12·0 (95% bootstrap CI 3·6–28·3) substantially exceeds the literature-derived prior
range of 2–5.[8,9] To test whether this discrepancy reflects a genuine feature of the data or a consequence of the model's reporting-fraction trade-off, we re-estimated all parameters with $\psi_{M\rightarrow N}$ fixed at 3·0 — the central value of the clinical literature prior — while allowing all other parameters, including both reporting fractions ($\rho_0$, $\rho_1$), to optimize freely.

#### Method

The gradient mask approach was used: the unconstrained parameter log($\psi_{M\rightarrow N}$) was pinned to log(3·0) at every optimization step, effectively removing its degree of freedom from the optimization. All 13 remaining free parameters were estimated by the same Adam optimizer with cosine decay schedule (2,000 steps, Learning Rate [LR] = 0·01) as the primary analysis.

#### Results

**Convergence and fit:**

| | **Primary (ψ free, Mode 1)** | **Fixed ψ = 3·0** | **Mode 2 (climate-ODE; global MLE)** |
|---|---|---|---|
| $\psi_{M\rightarrow N}$ | 12·0 | **3·00** (fixed) | ≈102 ($\rho_{mal}$ ≈ 81%) |
| Final negative log-likelihood (NLL) | 2,610 | **2,649** | 2,563 |
| ΔNLL vs. primary | — | **+39·5** | −46·9 |
| $\rho_0$ (malaria reporting) | 0·227 | **0·591** | ~0·81 |
| $\rho_1$ (iNTS reporting) | 0·020 | **0·033** | ~0·047 |
| $\beta_0$ (malaria base transmission) | 0·236 | 0·130 | — |

| | | | |
|---|---|---|---|
| $\beta_1$ (iNTS base transmission) | 0·085 | 0·110 | — |
| $\gamma_0$ (malaria recovery) | 0·00431 | 0·00399 | — |
| $\gamma_1$ (iNTS recovery) | 0·0127 | 0·00512 | — |
| $\kappa_0$ (malaria seasonality) | 0·308 | 0·272 | — |
| $\kappa_1$ (iNTS seasonality) | 0·246 | 0·457 | — |
| $r_0$ (malaria overdispersion) | 0·658 | 0·658 | — |
| $r_1$ (iNTS overdispersion) | 0·570 | 0·492 | — |

#### Interpretation

**The fixed-ψ model fits meaningfully worse. The ΔNLL of +39·5 over 2,000 training steps indicates that fixing** $\psi_{M\to N}$ = 3·0 reduces model fit compared to allowing it to optimize freely. Under standard likelihood ratio test criteria, a ΔNLL of 39·5 (equivalent to Δ−2logL = 79·0) for one constrained parameter represents a highly significant deterioration ($\chi^2_1$ critical value at p=0·001 is 10·8). The data therefore provide strong evidence that $\psi_{M\to N}$ > 3·0 when reporting fractions are simultaneously estimated.

**The reporting-fraction trade-off is confirmed. When** $\psi_{M\to N}$ is fixed at 3·0, the malaria reporting fraction jumps from 0·233 to 0·591. This confirms the structural non-identifiability described in S5: the model compensates for a lower susceptibility enhancement by requiring a much higher fraction of malaria cases to be observed (59% vs. 23%). A malaria passive surveillance ascertainment rate of 59% is plausible (facility-based RDT testing in high-transmission settings can achieve 40–70% capture of symptomatic cases), but is substantially higher than the 23% implied by Mode 1.

The iNTS reporting fraction also rises modestly (2·0% → 3·3%), though both values remain consistent with the extremely low ascertainment of NTS bacteremia in blood culture-based passive surveillance (typically 1–5% in sub-Saharan Africa).

**Other parameters shift coherently:**

- Malaria base transmission $\beta_0$ drops from 0·236 to 0·130 day$^{-1}$, consistent with the higher reporting fraction (fewer undetected infections required to explain observed counts)
- iNTS recovery rate $\gamma_1$ falls from 0·0127 to 0·0051 day$^{-1}$ (mean infectious period ~196 days vs. ~79 days), reflecting a longer carriage duration required to sustain iNTS counts under weaker malaria-mediated amplification
- iNTS seasonal amplitude $\kappa_1$ rises from 0·246 to 0·457, indicating stronger direct seasonal forcing (not mediated by malaria co-infection) in the fixed-ψ model

#### Conclusions from the sensitivity analysis

1. **The data prefer $\psi_{M\to N}$ > 3·0**: the fixed-ψ model fits significantly worse (ΔNLL = +39·5), providing evidence against the literature prior value as the true population-level susceptibility enhancement.
2. **The ρ/ψ trade-off is real and quantifiable: fixing** $\psi_{M\to N}$ = 3·0 requires malaria reporting to be 59% (vs. 23% in Mode 1). Both values are epidemiologically defensible,

but the discrepancy illustrates why external validation of $\rho_0$ or $\rho_1$ is needed to break the trade-off.

3. **The central qualitative conclusion is robust**: in all three solutions (Mode 1, Mode 2, fixed-ψ), $\psi_{M \to N}$ is substantially ≥ 3·0 (Mode 1: 12·0; Mode 2: ≈102 at $\rho_{mal}$ ≈ 81%; sensitivity lower bound: 3·0). All are consistent with the interpretation that active malaria materially increases NTS susceptibility, with a minimum plausible fold-change of approximately 3.
4. **Recommended next step**: a blood culture prevalence survey or record linkage with laboratory databases to obtain an independent estimate of $\rho_1$ would allow fixing this parameter externally, making $\psi_{M \to N}$ uniquely identifiable from the surveillance data.

### S12. Appendix References

Appendix references are numbered independently of the main manuscript reference list and are managed by EndNote.


1. Mossong J, Hens N, Jit M, et al. Social contacts and mixing patterns relevant to the spread of infectious diseases. PLoS Med. 2008;5(3):e74.

2. Hay SI, Rogers DJ, Toomer JF, Snow RW. Annual Plasmodium falciparum entomological inoculation rates across Africa. Trans R Soc Trop Med Hyg. 2000;94(2):113-127.

3. Aguas R, White LJ, Snow RW, Gomes MGM. Prospects for malaria eradication in sub-Saharan Africa. PLOS ONE. 2008;3(3):e1767.

4. Chitnis N, Hyman JM, Cushing JM. Determining important parameters in the spread of malaria through the sensitivity analysis of a mathematical model. Bull Math Biol. 2008;70(5):1272-1296.

5. Filipe JAN, Riley EM, Drakeley CJ, Sutherland CJ, Ghani AC. Determination of the processes driving the acquisition of immunity to malaria using a mathematical transmission model. PLOS Comput Biol. 2007;3(12):e255.

6. Yamba EI, Tompkins AM, Fink AH, et al. Climate drivers of malaria transmission seasonality in sub-Saharan Africa. GeoHealth. 2023;7(1):e2022GH000698.

7. Marks F, von Kalckreuth V, Aaby P, et al. Incidence of invasive Salmonella disease in sub-Saharan Africa: a multicentre population-based surveillance study. Lancet Glob Health. 2017;5(3):e310-e323.

8. Morpeth SC, Ramadhani HO, Crump JA. Invasive non-Typhi Salmonella disease in Africa. Clin Infect Dis. 2009;49(4):606-611.

9. Cunnington AJ, de Souza JB, Walther M, Riley EM. Malaria impairs resistance to Salmonella through heme- and heme oxygenase-dependent dysfunctional granulocyte mobilization. Nat Med. 2012;18(1):120-127.

10. Kingma DP, Ba J. Adam: A method for stochastic optimization. International Conference on Learning Representations (ICLR). 2015. arXiv:1412.6980.

11. VanderWeele TJ. Explanation in Causal Inference: Methods for Mediation and Interaction. Oxford University Press; 2015.

Sources listed in the original appendix bibliography that are not yet linked to a citation (see conversion notes; not EndNote-managed):

Eckhoff PA. Plasmodium falciparum infection durations and infectiousness are shaped by antigenic variation and immunity. PLOS ONE. 2012;7(9):e44950.

Griffin AJ, McSorley SJ. Development of protective immunity to Salmonella, a mucosal pathogen with a systemic agenda. Mucosal Immunol. 2011;4(4):371-382.

Guiraud I, Sow AG, Ba EH, et al. Population-based incidence, seasonality and serotype distribution of invasive salmonellosis in rural Burkina Faso. PLOS ONE. 2017;12(7):e0178577.

Mandal S, Sarkar RR, Sinha S. Mathematical models of malaria - a review. Malar J. 2011;10:202.

Rohringer A, Veneti L, Stuken A, et al. Risk factors associated with long-term shedding of non-typhoidal Salmonella in humans. Eur J Clin Microbiol Infect Dis. 2025.

Smith DL, McKenzie FE, Snow RW, Hay SI. Revisiting the basic reproductive number for malaria. PLOS Biol. 2007;5(3):e42.

Thindwa D, Chipeta MG, Henrion MYR, Gordon MA. Distinct climate influences on the risk of typhoid compared to invasive non-typhoid Salmonella disease in Blantyre, Malawi. Sci Rep. 2020;10:20310.

Uche IV, MacLennan CA, Saul A. A systematic review of incidence, risk factors and case fatality rates of iNTS disease in Africa (1966-2014). PLOS Negl Trop Dis. 2017;11(1):e0005118.

### Appendix Table S1. Interrupted Time Series Results

Full results from segmented negative binomial regression with Newey-West HAC-corrected standard errors, a 6-month (183-day) linear coverage ramp-up for both interventions, and z-scored weekly precipitation (NASA POWER MERRA2 PRECTOTCORR) as a climate covariate. Two breakpoints: PMC = perennial malaria chemoprophylaxis (sulfadoxine-pyrimethamine + amodiaquine), introduced November 1, 2023; R21/Matrix-M = recombinant circumsporozoite protein malaria vaccine, introduced October 1, 2024 (children <2 years). Level change ($\beta_2$) = incidence rate ratio at full coverage (end of ramp). Slope change ($\beta_3$) = per-day change in incidence trend after reaching full coverage. N = 443 weekly observations.

| Outcome | Breakpoint | Effect | IRR | 95% CI | p-value |
|---|---|---|---|---|---|
| Malaria | Nov 2023 (PMC) | Level change ($\beta_2$) | 0·197 | 0·096–0·401 | <0·001 |

| | | | | | |
|---|---|---|---|---|---|
| Malaria | Nov 2023 (PMC) | Slope change ($\beta_3$, per day) | 1·002 | 0·998–1·007 | 0·370 |
| Malaria | Oct 2024 (R21) | Level change ($\beta_2$) | 0·860 | 0·204–3·625 | 0·837 |
| Malaria | Oct 2024 (R21) | Slope change ($\beta_3$, per day) | 0·998 | 0·994–1·002 | 0·254 |
| iNTS | Nov 2023 (PMC) | Level change ($\beta_2$) | 0·972 | 0·418–2·262 | 0·948 |
| iNTS | Nov 2023 (PMC) | Slope change ($\beta_3$, per day) | 0·990 | 0·981–0·998 | 0·018 |
| iNTS | Oct 2024 (R21) | Level change ($\beta_2$) | 6·572 | 0·519–83·134 | 0·146 |
| iNTS | Oct 2024 (R21) | Slope change ($\beta_3$, per day) | 1·015 | 1·006–1·024 | 0·001 |

*Note:* An apparent iNTS level increase at the PMC breakpoint (IRR 2·766, 95% CI 1·453–5·267; p=0·002) arises only in a fully unadjusted segmented regression carrying no seasonal control of any kind, and is a confounding artifact of the coincident November rainy-season transition. It is removed by seasonal adjustment — principally the Fourier harmonics and the coverage-ramp specification — rather than by the precipitation covariate specifically: removing precipitation alone from the model above shifts the estimate only from 0·972 (95% CI 0·418–2·262; p=0·948) to 0·947 (95% CI 0·407–2·204; p=0·899). The full decomposition is given in appendix table S4. The key intervention signal is the significant declining iNTS trend post-ramp (IRR 0·990/day, 95% CI 0·981–0·998; p=0·018). The R21 level change for iNTS (IRR 6·572, 95% CI 0·519–83·1; p=0·146) is imprecise and reflects sparse post-R21 follow-up data rather than a true effect.

### Appendix Table S2. Mediation Analysis Results

Causal mediation analysis using parametric g-computation (VanderWeele[11] potential-outcomes framework). Primary lag: 14 days. Bootstrap resamples: 1,000. Note: Bootstrap CI for proportion mediated is numerically unstable due to sparse iNTS counts (N=507 total cases over 443 weeks) and TE near the null; the point estimate of ≈50% should be treated as hypothesis-generating.

| **Estimand** | **IRR** | **95% CI** |
|---|---|---|
| Total Effect (TE) | 0·846 | 0·48–1·40 |
| Natural Direct Effect (NDE) | 1·015 | 0·578–2·04 |
| Natural Indirect Effect (NIE) | 0·833 | 0·382–1·29 |
| Proportion Mediated (%) | 49·8 | −86·4 to >10^7 (unstable) |
| Population Attributable Fraction (%) | −29·5 | Numerically unstable |

### Appendix Table S3. ITS Coverage Ramp-Up Sensitivity Analysis

Results from segmented negative binomial ITS with Newey-West HAC correction, testing linear coverage ramp-up periods of 3, 4, 5, and 6 months for both PMC (Nov 2023) and R21/Matrix-M (Oct 2024) breakpoints simultaneously. All models include a z-scored weekly precipitation covariate (NASA POWER MERRA2 PRECTOTCORR) to control for rainfall-driven seasonal confounding. The 6-month ramp (183 days) was selected as the primary analysis. IRR = incidence rate ratio; CI = confidence interval.

**Malaria outcome**

| Ramp period | Breakpoint | Effect | IRR | 95% CI | p-value |
|---|---|---|---|---|---|
| 3 months (91 d) | Nov 2023 (PMC) | Level change | 0·311 | 0·156–0·620 | <0·001 |
| 3 months (91 d) | Nov 2023 (PMC) | Slope change/day | 0·999 | 0·996–1·002 | 0·540 |
| 3 months (91 d) | Oct 2024 (R21) | Level change | 1·322 | 0·563–3·103 | 0·521 |
| 3 months (91 d) | Oct 2024 (R21) | Slope change/day | 1·001 | 0·998–1·004 | 0·476 |
| 4 months (122 d) | Nov 2023 (PMC) | Level change | 0·265 | 0·131–0·536 | <0·001 |
| 4 months (122 d) | Nov 2023 (PMC) | Slope change/day | 1·000 | 0·996–1·003 | 0·963 |
| 4 months (122 d) | Oct 2024 (R21) | Level change | 1·198 | 0·430–3·340 | 0·730 |
| 4 months (122 d) | Oct 2024 (R21) | Slope change/day | 1·000 | 0·997–1·003 | 0·920 |
| 5 months (152 d) | Nov 2023 (PMC) | Level change | 0·225 | 0·112–0·451 | <0·001 |
| 5 months (152 d) | Nov 2023 (PMC) | Slope change/day | 1·001 | 0·997–1·005 | 0·587 |
| 5 months (152 d) | Oct 2024 (R21) | Level change | 0·993 | 0·296–3·335 | 0·991 |
| 5 months (152 d) | Oct 2024 (R21) | Slope change/day | 0·999 | 0·996–1·002 | 0·554 |
| 6 months (183 d) *(primary)* | Nov 2023 (PMC) | Level change | **0·197** | **0·096–0·401** | **<0·001** |
| 6 months (183 d) *(primary)* | Nov 2023 (PMC) | Slope change/day | **1·002** | **0·998–1·007** | **0·370** |
| 6 months (183 d) *(primary)* | Oct 2024 (R21) | Level change | **0·860** | **0·204–3·625** | **0·837** |
| 6 months (183 d) *(primary)* | Oct 2024 (R21) | Slope change/day | **0·998** | **0·994–1·002** | **0·254** |

**iNTS outcome**

| Ramp period | Breakpoint | Effect | IRR | 95% CI | p-value |
|---|---|---|---|---|---|
| 3 months (91 d) | Nov 2023 (PMC) | Level change | 1·014 | 0·438–2·347 | 0·975 |
| 3 months (91 d) | Nov 2023 (PMC) | Slope change/day | 0·996 | 0·990–1·001 | 0·146 |
| 3 months (91 d) | Oct 2024 (R21) | Level change | 0·593 | 0·118–2·969 | 0·525 |
| 3 months (91 d) | Oct 2024 (R21) | Slope change/day | 1·008 | 1·002–1·015 | 0·008 |
| 4 months (122 d) | Nov 2023 (PMC) | Level change | 0·904 | 0·385–2·125 | 0·818 |
| 4 months (122 d) | Nov 2023 (PMC) | Slope change/day | 0·996 | 0·989–1·002 | 0·194 |
| 4 months (122 d) | Oct 2024 (R21) | Level change | 0·642 | 0·093–4·435 | 0·653 |
| 4 months (122 d) | Oct 2024 (R21) | Slope change/day | 1·009 | 1·002–1·015 | 0·009 |
| 5 months (152 d) | Nov 2023 (PMC) | Level change | 0·902 | 0·378–2·151 | 0·816 |
| 5 months (152 d) | Nov 2023 (PMC) | Slope change/day | 0·994 | 0·987–1·001 | 0·086 |
| 5 months (152 d) | Oct 2024 (R21) | Level change | 1·264 | 0·168–9·496 | 0·820 |
| 5 months (152 d) | Oct 2024 (R21) | Slope change/day | 1·011 | 1·004–1·018 | 0·003 |
| 6 months (183 d) *(primary)* | Nov 2023 (PMC) | Level change | **0·972** | **0·418–2·262** | **0·948** |
| 6 months (183 d) *(primary)* | Nov 2023 (PMC) | Slope change/day | **0·990** | **0·981–0·998** | **0·018** |
| 6 months (183 d) *(primary)* | Oct 2024 (R21) | Level change | **6·572** | **0·519–83·1** | **0·146** |
| 6 months (183 d) *(primary)* | Oct 2024 (R21) | Slope change/day | **1·015** | **1·006–1·024** | **0·001** |

*Interpretation:* All models include a precipitation covariate (NASA POWER MERRA2 PRECTOTCORR, z-scored), which absorbs rainfall-driven seasonal confounding. Malaria PMC level changes are robust and consistent across all ramp periods (IRR range 0·197–0·311, all p<0·001), and are robust to removal of the precipitation covariate (appendix table S4). For iNTS, the PMC level change at rollout is null across all ramp periods when precipitation is controlled (IRR range 0·90–1·01, all p>0·8), the November iNTS increase seen in a fully unadjusted analysis was a seasonal confound coinciding with the rainy-season onset, and is removed by the Fourier harmonics and ramp specification rather than by the precipitation covariate specifically (appendix table S4). The iNTS declining slope post-PMC remains significant at the primary 6-month ramp (IRR 0·990/day, 95% CI 0·981–0·998; p=0·018), consistent with a lagged malaria-mediated effect. Bold rows indicate the primary analysis (6-month ramp).

### Appendix Table S4. ITS Specification Decomposition: Coverage Ramp, Fourier Seasonality, and Precipitation Adjustment

The primary ITS model (appendix table S1) differs from a fully unadjusted segmented regression in three respects: it models rollout as a 183-day linear coverage ramp rather than a binary step, it includes two Fourier harmonic pairs for seasonality, and it includes z-scored weekly precipitation. This table refits the model under all five nested specifications so that each contribution can be read separately. All specifications use Newey-West HAC standard errors (lag 10), a log(population) offset, and both breakpoints (PMC November 2023; R21/Matrix-M October 2024). N = 443 weekly observations throughout.

| Specification | Coverage term | Fourier harmonics | Precipitation covariate |
|---|---|---|---|
| **A (primary)** | **183-day linear ramp** | **2 pairs** | **Yes** |
| B | 183-day linear ramp | 2 pairs | No |
| C | Binary step | 2 pairs | Yes |
| D | Binary step | 2 pairs | No |
| **E (fully unadjusted)** | **Binary step** | **None** | **No** |

#### Malaria outcome

| Spec | Breakpoint | Effect | IRR | 95% CI | p-value |
|---|---|---|---|---|---|
| **A (primary)** | **Nov 2023 (PMC)** | **Level change (β2)** | **0·197** | **0·096–0·401** | **<0·001** |
| B | Nov 2023 (PMC) | Level change (β2) | 0·203 | 0·100–0·410 | <0·001 |
| C | Nov 2023 (PMC) | Level change (β2) | 0·590 | 0·324–1·075 | 0·085 |
| D | Nov 2023 (PMC) | Level change (β2) | 0·628 | 0·343–1·151 | 0·132 |
| E | Nov 2023 (PMC) | Level change (β2) | 0·943 | 0·516–1·723 | 0·848 |
| **A (primary)** | **Nov 2023 (PMC)** | **Slope change (β3, per day)** | **1·002** | **0·998–1·007** | **0·370** |
| B | Nov 2023 (PMC) | Slope change (β3, per day) | 1·002 | 0·997–1·006 | 0·386 |
| C | Nov 2023 (PMC) | Slope change (β3, per day) | 0·996 | 0·994–0·999 | 0·002 |
| D | Nov 2023 (PMC) | Slope change (β3, per day) | 0·996 | 0·994–0·998 | 0·002 |
| E | Nov 2023 (PMC) | Slope change (β3, per day) | 0·994 | 0·992–0·996 | <0·001 |
| **A (primary)** | **Oct 2024 (R21)** | **Level change (β2)** | **0·860** | **0·204–3·625** | **0·837** |
| B | Oct 2024 (R21) | Level change (β2) | 0·833 | 0·194–3·570 | 0·806 |
| C | Oct 2024 (R21) | Level change (β2) | 1·651 | 0·922–2·957 | 0·092 |
| D | Oct 2024 (R21) | Level change (β2) | 1·701 | 0·949–3·046 | 0·074 |

| | | | | | |
|---|---|---|---|---|---|
| E | Oct 2024 (R21) | Level change (β2) | 2·824 | 1·677–4·755 | <0·001 |
| **A (primary)** | **Oct 2024 (R21)** | **Slope change (β3, per day)** | **0·998** | **0·994–1·002** | **0·254** |
| B | Oct 2024 (R21) | Slope change (β3, per day) | 0·998 | 0·994–1·002 | 0·276 |
| C | Oct 2024 (R21) | Slope change (β3, per day) | 1·004 | 1·002–1·006 | <0·001 |
| D | Oct 2024 (R21) | Slope change (β3, per day) | 1·004 | 1·002–1·006 | <0·001 |
| E | Oct 2024 (R21) | Slope change (β3, per day) | 1·006 | 1·004–1·009 | <0·001 |

#### iNTS outcome

| Spec | Breakpoint | Effect | IRR | 95% CI | p-value |
|---|---|---|---|---|---|
| **A (primary)** | **Nov 2023 (PMC)** | **Level change (β2)** | **0·972** | **0·418–2·262** | **0·948** |
| B | Nov 2023 (PMC) | Level change (β2) | 0·947 | 0·407–2·204 | 0·899 |
| C | Nov 2023 (PMC) | Level change (β2) | 1·856 | 0·940–3·664 | 0·075 |
| D | Nov 2023 (PMC) | Level change (β2) | 1·754 | 0·881–3·493 | 0·110 |
| E | Nov 2023 (PMC) | Level change (β2) | 2·766 | 1·453–5·267 | 0·002 |
| **A (primary)** | **Nov 2023 (PMC)** | **Slope change (β3, per day)** | **0·990** | **0·981–0·998** | **0·018** |
| B | Nov 2023 (PMC) | Slope change (β3, per day) | 0·990 | 0·981–0·998 | 0·020 |
| C | Nov 2023 (PMC) | Slope change (β3, per day) | 0·995 | 0·991–0·998 | 0·006 |
| D | Nov 2023 (PMC) | Slope change (β3, per day) | 0·995 | 0·991–0·999 | 0·009 |
| E | Nov 2023 (PMC) | Slope change (β3, per day) | 0·993 | 0·989–0·996 | <0·001 |
| **A (primary)** | **Oct 2024 (R21)** | **Level change (β2)** | **6·572** | **0·519–83·134** | **0·146** |
| B | Oct 2024 (R21) | Level change (β2) | 6·449 | 0·508–81·908 | 0·151 |
| C | Oct 2024 (R21) | Level change (β2) | 0·439 | 0·119–1·613 | 0·215 |
| D | Oct 2024 (R21) | Level change (β2) | 0·426 | 0·116–1·557 | 0·197 |
| E | Oct 2024 (R21) | Level change (β2) | 0·520 | 0·168–1·609 | 0·256 |
| **A (primary)** | **Oct 2024 (R21)** | **Slope change (β3, per day)** | **1·015** | **1·006–1·024** | **0·001** |
| B | Oct 2024 (R21) | Slope change (β3, per day) | 1·015 | 1·006–1·024 | 0·001 |
| C | Oct 2024 (R21) | Slope change (β3, per day) | 1·009 | 1·004–1·014 | <0·001 |
| D | Oct 2024 (R21) | Slope change (β3, per day) | 1·009 | 1·004–1·014 | <0·001 |
| E | Oct 2024 (R21) | Slope change (β3, per day) | 1·012 | 1·008–1·016 | <0·001 |

**Model fit (log-likelihood)**

| Outcome | A | B | C | D | E |
|---|---|---|---|---|---|
| Malaria | −1896·5 | −1897·1 | −1897·2 | −1897·8 | −1905·5 |
| iNTS | −630·9 | −631·0 | −634·5 | −634·6 | −638·4 |

Interpretation: The precipitation covariate contributes little once Fourier harmonics are present. Comparing A with B — the only correct climate-adjusted versus climate-unadjusted contrast, since these differ solely in the precipitation term — the iNTS PMC level change moves from 0·972 (95% CI 0·418–2·262; p=0·948) to 0·947 (95% CI 0·407–2·204; p=0·899) and the malaria PMC level change from 0·197 (95% CI 0·096–0·401; p<0·001) to 0·203 (95% CI 0·100–0·410; p<0·001); no estimate changes materially, and the log-likelihood improves by less than 1 unit for either outcome. The large shifts in this table are driven instead by the coverage ramp (A→C, B→D) and by the Fourier harmonics (D→E). The apparent iNTS level increase at PMC rollout that motivated the seasonal-confounding interpretation appears only in specification E (IRR 2·766, 95% CI 1·453–5·267; p=0·002), which imposes no seasonal control of any kind; it is already absent once Fourier harmonics are added with no precipitation term at all (specification D, IRR 1·754, 95% CI 0·881–3·493; p=0·110). Specification E also produces the largest spurious malaria level change at the R21 breakpoint (IRR 2·824, 95% CI 1·677–4·755; p<0·001), an effect that is not plausibly attributable to a malaria vaccine and that likewise disappears under seasonal control. Specification E fits both outcomes worst on log-likelihood. The correct reading is therefore that seasonal control — principally the Fourier harmonics, together with the ramp specification — removes the confounding, and that the precipitation covariate is a marginal refinement rather than the decisive adjustment.

### Appendix Figure S1. Intervention Synergy Index

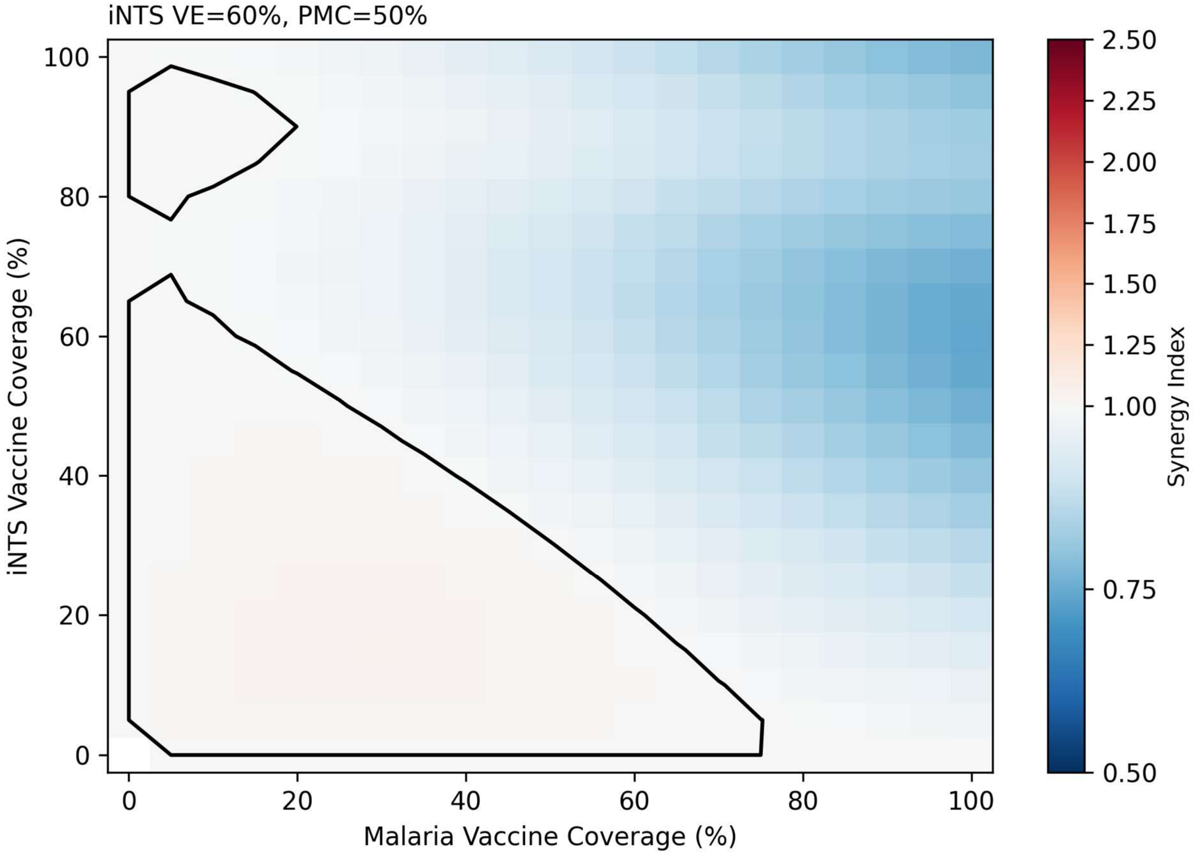


Synergy index heatmap showing the ratio of the combined intervention effect to the sum of individual effects, as a function of R21/Matrix-M coverage (x-axis) and iNTS vaccine coverage (y-axis), at 50% PMC coverage and 70% iNTS vaccine efficacy. Values >1 indicate super-additive (synergistic) effects; values <1 indicate sub-additivity. Black contour = SI = 1 (additivity boundary). Super-additivity arises from the malaria-NTS susceptibility interaction: reducing malaria amplifies the effectiveness of the iNTS vaccine by reducing the fraction of the population in an immunologically enhanced-susceptibility state.

### Appendix Figure S2. Univariate Marginal Effects

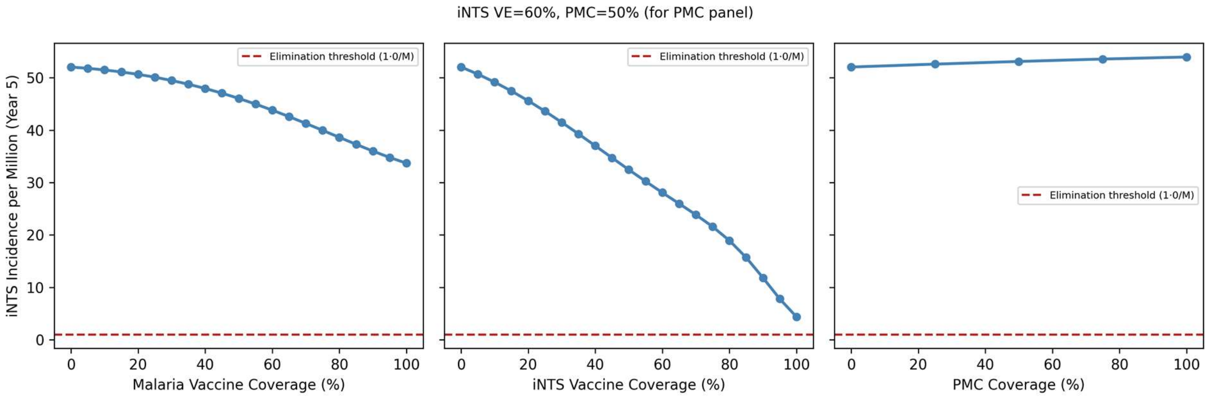


Year-5 NTS incidence per million as a function of each individual intervention at fixed baseline coverage for the other two. Left panel: R21/Matrix-M malaria vaccine coverage (0–100%); center panel: investigational iNTS vaccine coverage (0–100%); right panel: PMC coverage (0–100%). Red dashed line = elimination threshold (1 per million). PMC = perennial malaria chemoprophylaxis; R21/Matrix-M = recombinant circumsporozoite protein malaria vaccine.

### Appendix Figure S3. Elimination Boundary — NTS Incidence at Year 5

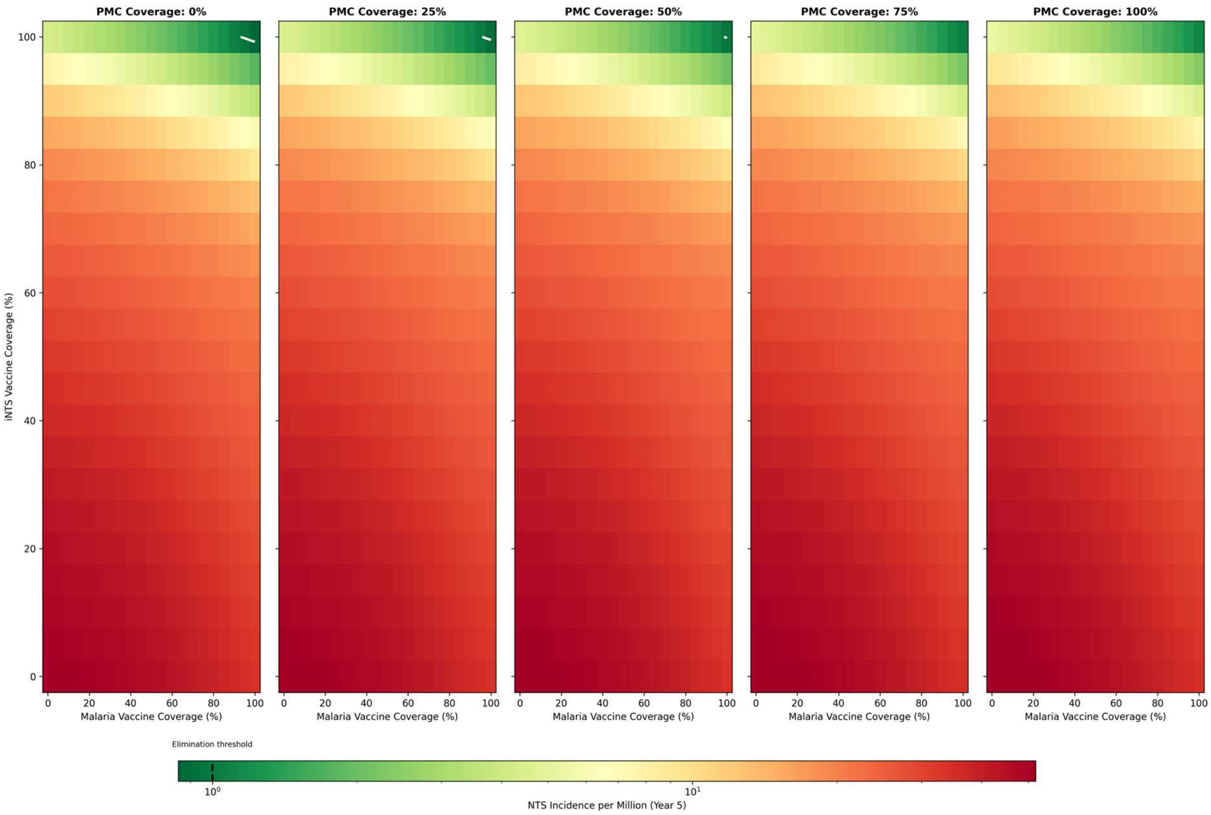


Five-panel heatmap showing projected year-5 NTS incidence per million (log-scale color gradient; green = near elimination, red = high burden) as a function of R21/Matrix-M malaria vaccine coverage (x-axis, 0–100%) and investigational iNTS vaccine coverage (y-axis, 0–100%), faceted by PMC coverage level (0%, 25%, 50%, 75%, 100%). Displayed at iNTS vaccine efficacy = 60%. White contour line = elimination threshold (1 per million). PMC = perennial malaria chemoprophylaxis (sulfadoxine-pyrimethamine + amodiaquine); R21/Matrix-M = recombinant circumsporozoite protein malaria vaccine. Results at other iNTS vaccine efficacy levels (30–90%) are available in the analysis data files.

### Appendix Figure S4. Causal Mediation Analysis

*Panel (a):*

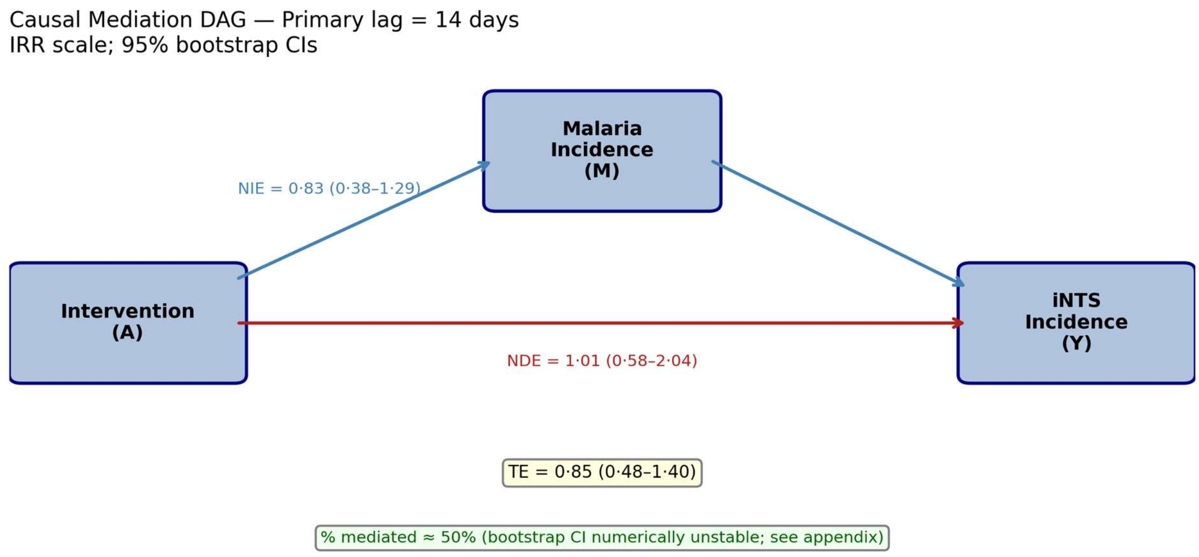


*Panel (b):*

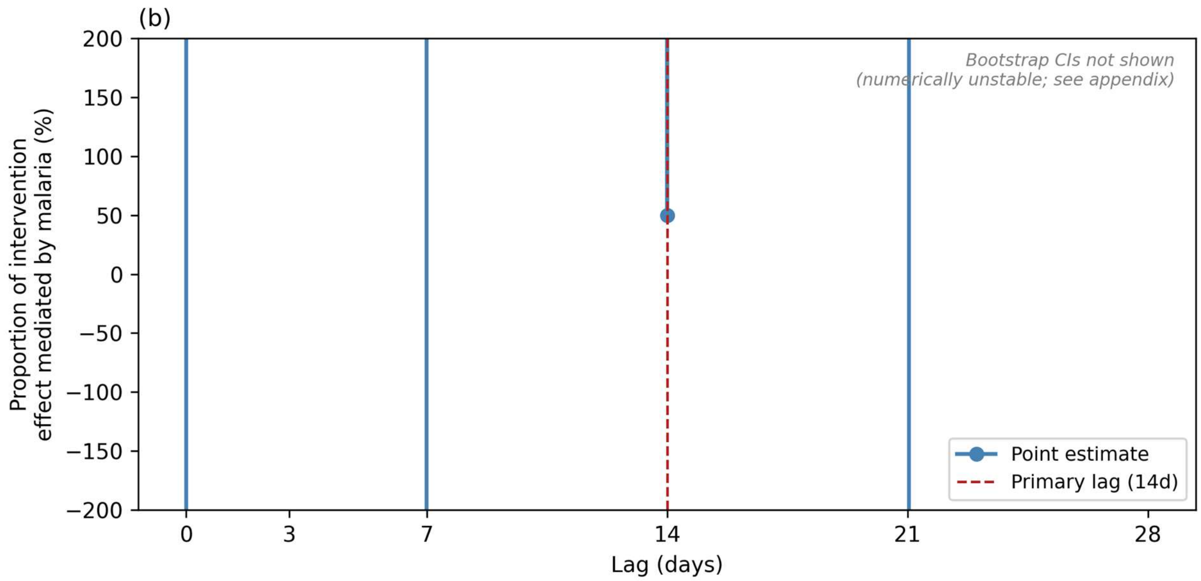


**(a)** Directed acyclic graph (DAG) of the causal mediation structure. The total effect of PMC on iNTS incidence is decomposed into a natural direct effect (NDE, red arrow; not operating through malaria) and a natural indirect effect (NIE, blue arrows; operating through first reducing malaria incidence). Effect estimates shown on the incidence rate ratio (IRR) scale at the primary 14-day lag (1,000 bootstrap resamples): TE = 0·846 (95% CI 0·48–1·40); NDE = 1·015 (0·578–2·04); NIE = 0·833 (0·382–1·29); proportion mediated ≈ 50% (bootstrap CI numerically unstable; see appendix S7).

**(b)** Proportion of total PMC effect on iNTS mediated through malaria as a function of assumed biological lag (0–28 days). The point estimate at the primary lag (14 days) is ≈50%; estimates at all other lags are numerically extreme (outside y-axis range). Bootstrap confidence intervals are not shown — they are numerically unstable due to sparse iNTS counts (507 cases over 443 weeks) and TE near the null; see appendix S7 for full explanation. Full numeric results are reported in appendix table S2.
